# The Online Health Safety Gap: Consensus Alignment Does Not Imply Safety in Peer-to-Peer Health Narratives

**DOI:** 10.64898/2026.09.18.26363448

**Authors:** Ommo Clark, Zeliatu Ahmed, Karuna P. Joshi

## Abstract

Online health information systems evaluate content by its alignment with medical consensus, treating that alignment as a reliable signal of safety. In peer-to-peer health discourse, that assumption fails: advice that is factually accurate is not necessarily safe to act on. This paper identifies and empirically validates the **Online Health Safety Gap**, a structural misspecification in which medically accurate content can carry substantial health risk. Across 704 health narratives (699 classified), two scores computed from non-overlapping feature sets, Narrative Truth Distance for epistemic divergence and Narrative Risk Score for health risk potential, share only 4.9% of their variance (*r* = 0.222, *p* < 0.001). Consequently, 39.6% of narratives fall in the two off-diagonal quadrants that single-axis systems mishandle by construction: aligned-but- risky content (25.2%) invisible to fact-checking, and divergent-but-safe content (14.4%) incorrectly suppressed. The same independence constrains what existing benchmarks can measure. On an expert-labeled misinformation benchmark of 437 posts containing 127 misinformation instances, the highest discrimination attained is Youden’s *J* of 0.349, by a supervised classifier trained on those labels directly, which neither frozen biomedical embeddings nor a prompted large language model improves upon. Labels encoding factual accuracy therefore carry little signal about behavioral risk, and benchmarks of this construction cannot evaluate risk-aware assessment. To operationalize the gap, we introduce the **Classification Quadrant**, which maps epistemic divergence and health risk potential onto four governance categories with distinct intervention implications, and argue for a shift from fact- centric evaluation to risk-aware assessment.

## 1 Introduction

Consider this Reddit post: “I’ve been on metformin 500 mg twice daily but my fasting glucose is still 160-180 mg/dL. I researched the standard dose and increased it to 1000 mg myself. Two weeks later my glucose is 95-110. Finally under control!” Every clinical detail is accurate. Metformin is a first-line treatment for type 2 diabetes, the dosages fall within standard prescribing ranges, and the reported glucose improvement is physiologically plausible [1]. A fact-checking system will find no disagreement with medical consensus. Yet the narrative describes unsupervised dose escalation, a behavior that carries genuine health risk when undertaken without medical assessment of renal function, medication interactions, or clinical appropriateness for the individual’s specific circumstances. While dose titration can be a clinician-taught self-management skill in chronic disease care, the narrative provides no indication of medical oversight, and the risk lies precisely in this absence rather than in the act of adjustment itself. This harm potential is invisible to fact-checking and misinformation detection systems, which operate on a single axis of epistemic alignment and evaluate facts and keywords rather than the behavior a narrative describes. The failure is one of scope rather than of accuracy. We term it the **Online Health Safety Gap**: the structural inability of fact-centric systems to detect harmful behaviors embedded in narratives.

Individuals increasingly turn to peer-to-peer online health content to make consequential self-care decisions such as adjusting medications, discontinuing treatments, and delaying professional consultation. A prior qualitative study across seven countries found that 87% of respondents regularly used online health information for symptom checking, self-diagnosis, and treatment; however, despite concerns about its accuracy, they continued to rely on it to inform health decisions, driven by convenience, cost, and barriers to access [2], [3]. This highlights a critical gap in the information environment, where no reliable mechanism exists to determine whether a given narrative, however credible it appears, is safe to act on.

We define *Online Health Safety* as the protection of individuals from harm arising from self-care decisions guided by online health information in non-clinical settings. This problem space is distinct from patient safety [4], which assumes clinician involvement and institutional oversight, and from general misinformation detection [5], [6], which treats factual incorrectness as the primary harm vector. The Online Health Safety Gap opens between these paradigms precisely because in peer-to-peer self-care settings, the primary harm vector is not factual error but the contextually inappropriate application of accurate information.

This paper makes three contributions. The first and principal contribution is the identification and empirical validation of the Online Health Safety Gap through three converging bodies of evidence. A formal independence analysis establishes that epistemic divergence and health risk potential are substantially non-redundant, sharing 4.9% of their variance (Section 6.2). A prevalence analysis shows that 25.2% of narratives in the studied corpus are factually aligned with established medical consensus yet carry elevated health risk, and are therefore approved by any system that evaluates alignment alone (Section 6.3). An evaluation on an expert-labeled benchmark establishes that the separability of binary misinformation labels is bounded at Youden’s *J* = 0.349 (95% CI [0.252,0.447]) by a supervised classifier trained on those labels directly, so benchmarks of this construction cannot evaluate risk-aware assessment (Section 6.4).

Together these establish that medical fluency is not a proxy for safety: how far a narrative departs from consensus says little about whether acting on it would cause harm. The second contribution is the **Classification Quadrant**, a two-dimensional governance structure that partitions the credibility–risk space into four distinct categories with proportionate intervention implications (Section 3.3). The third contribution is the **formalization of online health safety as a distinct problem space** (Section 3.1), providing the conceptual foundation from which benchmark design, content moderation architecture, and AI guardrail objectives can be systematically reassessed.

Two research questions frame the evaluation:

- **RQ1:** What is the prevalence of aligned-but-risky and divergent-but-safe narratives in peer-to-peer health discourse, and what share of the corpus do single-axis systems mishandle by construction?
- **RQ2:** What evaluation signal do existing expert-labeled misinformation benchmarks provide for risk-aware assessment, and what does the answer imply for benchmark design?

The paper is organized as follows. Section 2 reviews related work. Section 3 defines online health safety and introduces the Classification Quadrant. Section 4 summarizes the VERITAS (**V**erification **E**ngine for **R**isk-aware **I**nformation **T**rust **A**ssessment in health **S**tories) pipeline that produces the NTD and NRS scores. Section 5 describes the experimental design. Section 6 presents empirical results. Section 7 discusses implications. Section 8 concludes.

## 2 Related Work

Addressing online health safety requires engagement with four bodies of research: computational health misinformation detection, theoretical frameworks for risk and narrative structure, knowledge-grounded and neuro-symbolic methods, and content governance research.

### 2.1 Fact-Checking and Misinformation Detection

Computational research on health misinformation has developed along three principal lines: claim detection and verification [7], stance and propagation-aware classification [8], [9], and knowledge-grounded verification using biomedical resources [10], [11], [12]. Systems such as ClaimBuster [7] extract propositional claims and match them against authoritative sources, while Biomedical Bidirectional Encoder Representations from Transformers (BioBERT)-based architectures [13] have improved medical entity extraction and enabled fact-checking grounded in structured resources including the Unified Medical Language System (UMLS) [14], the Systematized Nomenclature of Medicine Clinical Terms (SNOMED CT) [15], and the Semantic MEDLINE Database (SemMedDB) [16]. Across these approaches the output is a single label indicating whether claims agree with medical consensus.

Recall for safety-critical content has been consistently low across architectural generations. Baseline natural language processing (NLP) classifiers achieve as little as 12% recall for health misinformation [17], GPT-4o reaches only 34% recall for underrepresented mental health categories despite high aggregate accuracy [18], and a systematic review of 33 cancer-information extraction studies reports F1-scores as low as 0.355 [19]. The consistency of these deficits across otherwise heterogeneous systems points to a shared structural cause rather than model-specific shortcomings. A further challenge is the asymmetric treatment of misclassification cost; binary NLP systems assume symmetric error costs, yet failing to identify potentially harmful content carries fundamentally different consequences from incorrectly flagging benign content [20]. The binary output structure offers no graduated pathway between approval and rejection.

The evaluation infrastructure shares the same single-axis objective. Health misinformation corpora annotate content for correctness relative to medical consensus [10], [21], so systems are measured on the dimension they were optimized for, and behavioral risk enters neither the objective nor the measurement. Whether labels of this construction can separate risk-aware assessment from divergence-based assessment is an empirical question, examined in Section 6.4.

### 2.2 Risk, Narrative Theory, and Causal Reasoning

Risk analysis research offers formalisms that the misinformation detection literature has not absorbed. The Kaplan-Garrick framework [22] decomposes risk into scenario identification, probability estimation, and consequence assessment, three evaluative tasks that current systems reduce to one question of truthfulness. Aven [23] further emphasized uncertainty in consequence assessment, a concern directly relevant to health narratives where outcomes depend on individual clinical context that online content cannot specify.

Narrative theory supplies a parallel foundation. Labov and Waletzky’s sociolinguistic framework [24] characterizes personal experience narratives as coherent sequences with identifiable stages rather than as collections of isolated claims. Representing online health discussion at the claim level discards the temporal, agentive, and consequential structure that distinguishes a supervised from an unsupervised course of self-medication. Ranade et al. [25] identified five critical gaps in computational narrative understanding, including the inability to reason over narrative structure and the failure to assess consequences of described actions, that directly motivate narrative-aware assessment. Prior qualitative work [2], [3] further shows that individuals exercise context-dependent, narrative-sensitive trust assessments when engaging with online health information, confirming that verification confined to propositional correctness captures only part of what users implicitly evaluate.

Pearl’s causal hierarchy [26] provides the final theoretical strand, distinguishing between what is, what would happen if an action were taken, and what would have happened under different conditions. Assessing whether a described behavior is safe to act on requires interventional and counterfactual reasoning, not merely observational claim-matching. Recent causal NLP advances [27], [28] have developed text-level causal reasoning methods, but these have not been applied to independent credibility and risk assessment over health narrative structures.

### 2.3 Knowledge-Grounded and Neuro-Symbolic Approaches

Knowledge-graph-enhanced verification grounds claims in structured biomedical resources and validates entity-level relationships [29]. This reliably captures propositional correctness but cannot assess whether a described sequence of actions such as self-adjusting a dose, monitoring personal outcomes, or deferring consultation, constitutes safe behavior. Knowledge graphs encode facts about entities and their relationships, not judgments about the contextual appropriateness of acting on those facts.

Neuro-symbolic integration [30], [31], [32] combines neural pattern recognition with symbolic reasoning, with recent biomedical applications including explainable diagnosis prediction [33] and knowledge-graph-enhanced question answering [34]. This paradigm is well suited to problems requiring both pattern recognition over unstructured text and principled reasoning over structured knowledge, precisely the combination that online health safety assessment demands. The VERITAS framework [35], [36], developed in our prior work and summarized in Section 4, applies neuro-symbolic methods to compute two separate continuous scores, Narrative Truth Distance (NTD) and Narrative Risk Score (NRS), over structured health narrative representations. The present paper establishes the gap these scores jointly reveal and introduces the Classification Quadrant that makes it actionable.

### 2.4 Content Governance and AI Guardrails

Content moderation research has established that proportionate, graduated interventions outperform binary accept/reject regimes in balancing informational access against harm reduction [20], [37], [38]. The EU AI Act [39] codifies this principle in regulatory form, requiring that intervention severity be proportionate to assessed risk. Operationalizing proportionality, however, requires an assessment infrastructure capable of distinguishing low-risk divergence from high-risk alignment, a discrimination that fact-checking cannot compute.

A parallel concern has emerged around large language models (LLMs), which increasingly mediate health information through chat interfaces and retrieval-augmented assistants [40], [41]. Guardrail systems focused on preventing hallucinations [42] inherit the same epistemic-only objective that limits binary classifiers; when an LLM produces consensus-consistent output encoding an unsafe action pattern, hallucination-focused guardrails do not intervene because no factual error has occurred.

### 2.5 The Gap the Literature Leaves

The four bodies of research reviewed above converge on the same unfilled space. Risk, narrative, and causal frameworks supply the theoretical vocabulary for two-dimensional assessment but have not been integrated into a classification structure applicable to peer-to-peer health discussion outside clinical settings. Neuro-symbolic methods enable the independent scoring such a structure requires, and governance research demands the graduated interventions it would support, yet no existing framework jointly assesses credibility and health risk potential and translates that joint assessment into actionable categories.

The closest prior work is Sehat et al. [20], which argues that fact-checking should be prioritized by potential harm rather than by falsity alone, and supplies a structured vocabulary for doing so. That proposal treats harm as a criterion for ranking content already identified as false, so harm modulates the response to detected divergence rather than being assessed on a pathway of its own. Content that is not false is therefore never ranked, and the aligned-but-risky case remains outside the scope of the prioritization. The step this paper takes is to compute harm potential independently of divergence, so that content raising no epistemic flag can still be surfaced.

The absence also extends to how the problem space is bounded. Patient safety research addresses harm from health decisions but presumes clinician involvement, institutional protocol, and regulated medication pathways [43], [44], none of which holds when individuals act on peer narratives without professional contact [45]. Misinformation research addresses unmediated public information but takes factual incorrectness as the harm vector. Neither literature formalizes the setting in which consequential self-care decisions are made outside clinical oversight and the harm arises from the application of accurate information, which is the problem space Section 3.1 defines.

The absence extends finally to evaluation: with corpora annotated for correctness alone, a system that assessed harm independently could not be distinguished from one that did not. The Classification Quadrant, introduced in the next section, fills this space.

## 3 The Online Health Safety Gap and the Classification Quadrant

### 3.1 Online Health Safety as a Distinct Problem Space

Online health safety, defined here as the protection of individuals from harm arising from self-care decisions guided by online health information in non-clinical settings, occupies a problem space distinct from its two nearest neighbors. Patient safety frameworks [43], [44] assume clinician involvement, institutional protocols, and regulated medication pathways, assumptions that do not hold when individuals self-diagnose or medicate based on peer-to-peer online advice [45]. General misinformation detection [8], [10], [21] assumes factual incorrectness is the primary harm vector, yet content that aligns with medical consensus can carry substantial health risk when acted on without medical supervision. Online health safety sits between these paradigms: it inherits from patient safety the concern with behavioral consequence, and from misinformation research the setting of unmediated public information, while sharing neither the institutional controls of the former nor the epistemic premise of the latter.

An important distinction within this problem space concerns clinician-taught self-management skills versus unsupervised self-medication. In chronic disease care, clinicians routinely teach patients to titrate medication doses within prescribed parameters (e.g., insulin sliding-scale adjustments, antihypertensive dose stepping) as part of structured self-management plans [45]. These clinician-sanctioned adjustments are a feature of successful patient education, not a safety failure. Online health safety concerns a different phenomenon: behaviors undertaken without any clinical framework, where the individual’s decision to adjust, discontinue, or combine treatments is guided by information obtained from peer narratives rather than professional guidance. The VERITAS pipeline’s NRS component treats the absence of clinical oversight indicators (consultation references, disclaimer language, professional guidance cues) as a risk signal; the act of self-management itself is not penalized.

### 3.2 Why Two Dimensions Are Required

Online health safety assessment operates along two axes: epistemic divergence *D*(*N*), measuring how far a narrative’s claims depart from established medical consensus, and health risk potential *R*(*N*), quantifying the harm potential if the described behavior is acted on. How strongly these axes covary determines whether the dominant failure mode in peer-to-peer health discourse is detectable at all. To the extent that *D*(*N*) predicts *R*(*N*), a system measuring divergence recovers risk implicitly and one-dimensional assessment suffices. To the extent that it does not, the residual is information that no divergence measurement can reach, however well calibrated.

The measured association falls decisively toward the second case. Seven convergent statistical tests place it below the pre-specified practical-independence criterion (Section 6.2), with 4.9% shared variance, so 95.1% of the variance in health risk potential lies outside what divergence explains. The mechanistic account is direct. Medically fluent language is equally available for describing safe and unsafe behaviors. The correct drug names, plausible dosages, and appropriate clinical measurements that signal credibility to a fact-checking system are precisely the vocabulary readily available to individuals [2]. A post describing unsupervised insulin dose adjustment uses identical vocabulary to one describing physician-supervised adjustment; the risk lies in the implied behavior and the absence of clinical oversight, neither of which is visible to lexical or entity-level verification.

Fact-checking and misinformation detection systems approximate correctness as a function of divergence alone:

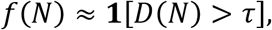

projecting the two-dimensional assessment space onto the *D*-axis and producing a single label. This projection discards health risk information entirely. The resulting mishandling is not incidental but the systematic consequence of the projection: content aligned with consensus but behaviorally risky is approved, and content diverging from consensus but behaviorally benign is suppressed. The two-dimensional assessment space must therefore be preserved rather than projected.

### 3.3 The Classification Quadrant

Discretizing each axis into “low” and “high” regions yields four categories, each corresponding to a distinct combination of epistemic divergence and health risk potential. We term this partition the **Classification Quadrant**. It assigns each narrative to one of four categories based on the threshold pair (*τ*_NTD_, *τ*_NRS_) applied to its NTD and NRS scores, which are computed from non-overlapping feature sets, making the joint credibility–risk profile visible, explainable, and actionable. Figure 1 presents the partition.

**Figure 1.**
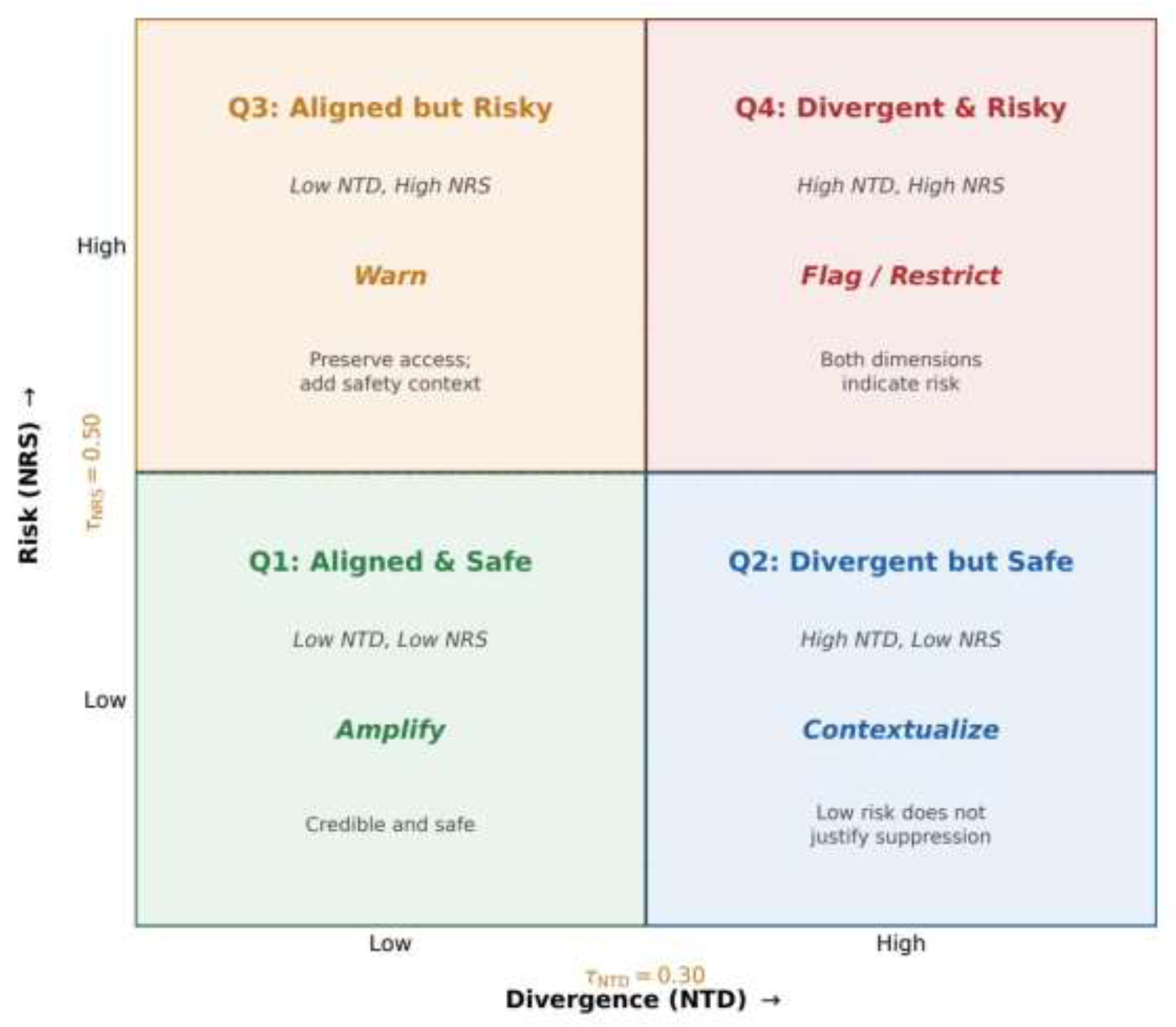
The Classification Quadrant. The NTD–NRS plane is partitioned by operational thresholds τ_NTD_ = 0.30 (vertical) and τ_NRS_ = 0.50 (horizontal) into four categories, each annotated with its threshold-based criteria, the proportionate governance intervention, and the rationale linking intervention to evidence. Interventions follow the graduated-response principle: amplify for Q1, contextualize for Q2, warn for Q3, and flag/restrict for Q4. Fact-checking and misinformation detection systems project this plane onto the D-axis alone (Equation [eq:binary_projection]), producing one label and discarding all health risk information. The Quadrant preserves both dimensions as independently actionable.

The four categories are defined by their joint credibility–risk position:

- **Q1: Aligned & Safe** (NTD < *τ*_NTD_ ∧ NRS < *τ*_NRS_). Narratives consistent with medical consensus describing behaviors with low harm potential.
- **Q2: Divergent but Safe** (NTD ≥ *τ*_NTD_ ∧ NRS < *τ*_NRS_). Narratives diverging from biomedical consensus but implying actions with minimal harm, including traditional and culturally grounded practices that depart from biomedical orthodoxy without posing substantive health risk.
- **Q3: Aligned but Risky** (NTD < *τ*_NTD_ ∧ NRS ≥ *τ*_NRS_). Narratives aligned with consensus describing behaviors such as self-directed medication adjustment, abrupt treatment discontinuation, and deferral of professional consultation, with significant harm potential if acted on without clinical supervision. Binary verification approves such narratives because its detection trigger, epistemic mismatch, is absent.
- **Q4: Divergent & Risky** (NTD ≥ *τ*_NTD_ ∧ NRS ≥ *τ*_NRS_). Narratives that both diverge from consensus and carry high harm potential, the category that fact-checking and misinformation detection systems are designed to detect.

Q1 and Q4 are categories in which epistemic alignment and harm status co-vary, and fact-checking and misinformation detection systems produce the correct outcome for them by coincidence. Q2 and Q3 are the categories in which the two dimensions diverge, making single-axis mishandling a structural necessity rather than an empirical shortcoming. Figure 4 illustrates each quadrant with a representative worked example. Whether Q2 and Q3 occur at rates sufficient to render claim-level and keyword-level verification inadequate is the empirical question addressed in Section 6.3.

#### 3.3.1 Governance implications

The four categories map to distinct, proportionate governance responses presented in Figure 1, following the graduated response principle established in content moderation research [20], [37], [38] and the proportionality criterion in the EU AI Act [39].

Q1 content warrants amplification because reliable first-person accounts of safe self-management inform others engaged in similar decisions [38]. Q2 content warrants contextual annotation rather than suppression because low health risk does not justify restricting informational access; this category also carries equity implications, as divergence-based suppression applied to Q2 disproportionately affects culturally grounded health practices that are safe but terminologically distant from Western biomedical consensus [46]. Q3 content warrants targeted safety prompts accompanying rather than replacing the content, since these narratives are consensus-aligned and removal sacrifices genuine informational value, while the harm is conditional on action and a warning addresses that conditional directly [47], [48]. Q4 content warrants the strongest intervention because both dimensions independently indicate risk [49].

The four-category structure is the minimum partition that preserves both dimensions as independently actionable while mapping each joint position to a distinct governance response. Collapsing to three categories would conflate Q2 with Q3, eliminating the distinction the Quadrant exists to make visible.

The thresholds *τ*_NTD_ and *τ*_NRS_ are operational decision boundaries, not claims that risk changes categorically at a particular value. Risk in the medical domain is a spectrum, and the NRS represents it as one: the score is a continuous sigmoid whose centering constant is calibrated so that the midpoint of the raw risk distribution maps to NRS = 0.50 (Appendix B). The threshold therefore marks the median of a graded scale rather than a categorical boundary, and the continuous scores are preserved throughout the pipeline and remain available for gradient-based deployment. In the studied corpus, 19.7% of narratives (138/699) fall within ±0.05 of that midpoint (NRS ∈ [0.45,0.55]), constituting a transition zone where classification confidence is lower. For narratives in this region, graduated responses such as softer advisory language or tiered confidence indicators are more appropriate than a binary warn or no-warn decision. The weight sensitivity analysis in Section 6.3 confirms that the core findings are robust across NTD pillar weightings, consistent with thresholds that are operational rather than fundamental.

### 3.4 Narrative-Level Representation

The risk features distinguishing aligned-and-safe from aligned-but-risky content, whether dose adjustment is supervised, whether treatment discontinuation is planned or abrupt, whether consultation was sought or deferred, are properties of narrative structure representing the agent-action-outcome relationships unfolding across a coherent sequence [24], [25]. A claim-level system can verify that “metformin treats type 2 diabetes” but cannot assess whether self- escalating the dose without medical review constitutes safe behavior. Any implementation of the Classification Quadrant therefore requires upstream representation preserving action sequences, agent roles, and outcome relationships, with architectural decoupling of epistemic and harm assessment so that neither pathway contaminates the other. VERITAS, described in Section 4, provides this infrastructure.

## 4 The VERITAS Framework

The Classification Quadrant operates on two separately computed scores produced by the VERITAS (**V**erification **E**ngine for **R**isk-aware **I**nformation **T**rust **A**ssessment in health **S**tories) pipeline, developed and validated in prior work [35], [36]. The architectural commitment enabling two-dimensional assessment is the strict separation of the NTD and NRS scoring pathways: both are computed from non-overlapping feature sets (ℱ_NTD_ ∩ ℱ_NRS_ = Ø) over the same structured narrative graph, so that no feature influences both scores and neither pathway can contaminate the other. Figure 2 presents the three-phase architecture.

**Figure 2.**
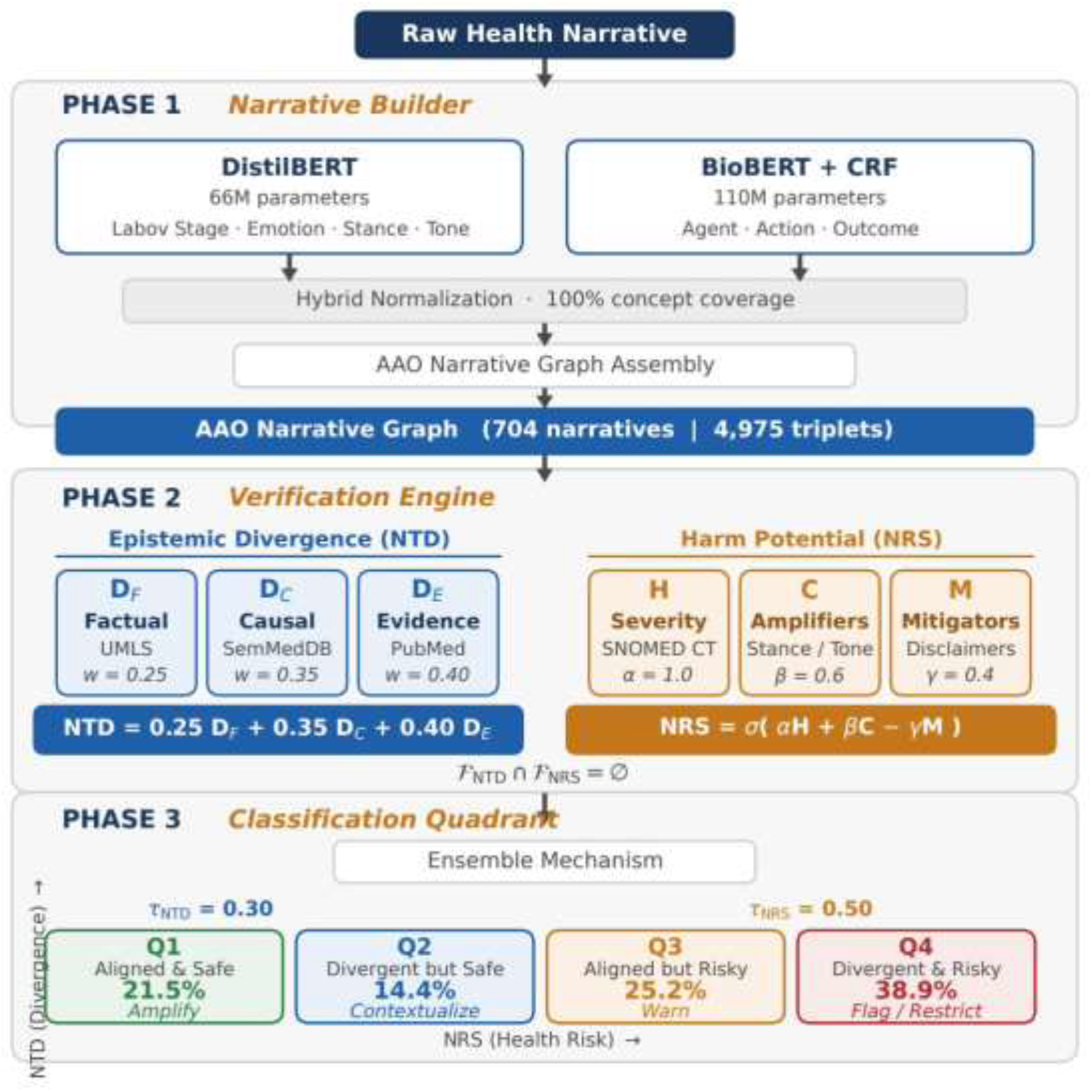
VERITAS neuro-symbolic three-phase pipeline. Phase 1 constructs Agent-Action-Outcome (AAO) narrative graphs from unstructured peer-to-peer health posts. Phase 2 computes NTD and NRS from non-overlapping feature sets, so that neither score can be recovered from the other’s inputs. Phase 3, the Classification Quadrant, maps the joint NTD–NRS position to one of four categories with graduated governance implications.

Phase 1, the *Narrative Builder* [35], reconstructs unstructured posts into structured Agent-Action-Outcome (AAO) narrative graphs, assigning Labovian narrative stages alongside emotion, stance, and tone labels so that computational systems can read posts as whole stories rather than isolated facts or keywords. The reconstruction employs two transformer models: a fine-tuned DistilBERT (66M parameters) for multi-task classification of Labovian stage, emotion, stance, and tone (per-task macro-F1 = 0.501 for Labovian stage, 0.466 emotion, 0.528 stance, and 0.465 tone under five-fold grouped cross-validation), and a BioBERT+CRF pipeline (110M parameters) for AAO entity span extraction (held-out span-F1 micro = 0.846, macro = 0.831; cross-validated span-F1 = 0.851 ± 0.007). Appendix C reports the full Phase 1 architecture, data partitioning protocol, and validation results, including per-entity-type extraction performance and baseline comparisons, so that this component can be assessed independently of [35]. Extracted entities undergo hybrid normalization against UMLS [14], achieving 100% concept coverage through a multi-strategy pipeline combining exact match, approximate string matching, and contextual disambiguation. The resulting narrative graph is the shared substrate from which both scoring pathways draw their respective, disjoint features. Phase 2a computes NTD through three verification pillars:

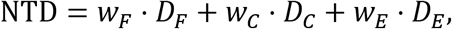

where *D_F_* measures concept-level alignment against UMLS [14], *D_C_* validates causal claims against SemMedDB [16], and *D_E_* assesses confidence calibration against PubMed, with weights *w_F_* = 0.25, *w_C_* = 0.35, *w_E_* = 0.40 derived in Appendix A. In plain terms, the NTD asks how far a narrative’s claims depart from what evidence-based medicine says. Phase 2b computes the NRS through an extended Kaplan-Garrick formalism [22]:

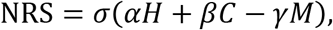

where *H* combines SNOMED CT [15] clinical severity, temporal immediacy, and user vulnerability; *C* captures contextual amplifiers including certainty mismatch and emotional intensity; and *M* represents protective mitigators such as disclaimers and references to professional consultation, with parameters *α* = 1.0, *β* = 0.6, *γ* = 0.4. In plain terms, the NRS asks how much harm could follow if a reader acts on the behavior a narrative describes. The parameters are fixed a priori and are not fitted to any label set; Appendix B specifies the sub- components, their weights, the basis on which each was set, and the scope of the validation claimed for the score. Because the two pathways read disjoint features, the residual association between the scores reported in Section 6.2 is a property of the corpus rather than of shared inputs.

### 4.1 Phase 3: Quadrant Assignment and Explanation

Phase 3 receives the NTD and NRS scores and applies the classification function:

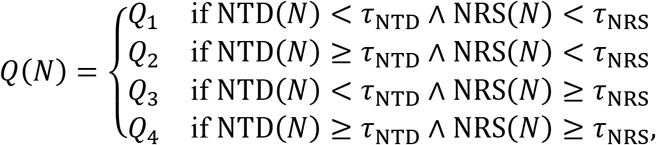

with operational thresholds *τ*_NTD_ = 0.30 and *τ*_NRS_ = 0.50. The risk threshold follows from the NRS construction: the centering constant maps a raw risk of 0.75, the midpoint of the raw-risk scale, to NRS = 0.50, so the threshold is a fixed point of the transformation rather than a categorical boundary. It is not the median of the empirical score distribution. The raw risk median on the classified corpus is 0.790, so the midpoint sits just below the median and *τ*_NRS_ = 0.50 falls at approximately the 36th percentile of the NRS distribution (median 0.560), with 448 of 699 narratives (64.1%) above it (Appendix B). The divergence threshold was set by expert review and verified for robustness across NTD pillar weightings (Section 6.3). Both were fixed on Corpus 1 before any evaluation, and neither is tuned against outcome labels.

Alongside each quadrant assignment, Phase 3 generates simple-language explanations decomposing the classification into its constituent signals. For a narrative in Q3, for example, the explanation identifies the specific action–outcome patterns driving the NRS score (“unsupervised dose escalation from 500 mg to 1000 mg”) while separately noting the basis for low divergence (“correct medication name, plausible dosage range, physiologically consistent glucose response”). In the non-clinical, peer-to-peer environment, there is no clinician to interpret the output on the user’s behalf; the explanation is therefore a structural requirement, not an optional feature. Without it, a categorical label communicates a verdict but not the reasoning that allows a user to determine whether the assessed risk applies to their own circumstances.

## 5 Experimental Design

### 5.1 Datasets and Annotation

Both corpora are drawn from a single collection of 5,386 threads spanning February 2019 to November 2024, gathered via the Python Reddit API Wrapper (PRAW v7.6.0) in compliance with Reddit’s Terms of Service and established ethical guidelines for internet research [50]. Collection used two strategies. First, 2,000 high-engagement threads were retrieved by keyword search across r/AskDocs, r/Coronavirus, r/COVID19, r/health, r/medicine, r/nutrition, and r/science. Second, 1,185 threads were collected directly from r/diabetes, r/fitness, r/health, r/hypertension, and r/nutrition. A further 2,201 threads with extended narrative content were drawn from the same health-focused sources; subreddit-level provenance was not retained for this batch (2,000 + 1,185 + 2,201 = 5,386). The health topics represented correspond to those most commonly searched in prior qualitative studies [2], [3]. No usernames or personally identifiable information were collected; examples are paraphrased to prevent reverse identification.

The two corpora were constructed independently, at different times, for different purposes, and under different annotation protocols. They also differ in the textual unit they comprise, and that difference governs what each can be used to establish.

#### 5.1.1 Corpus 1: Quadrant-annotated corpus (*N* = **704)**

From the master collection, 2,000 narrative segments meeting minimum inclusion criteria (≥50 words, personal health experience, identifiable temporal sequence) were annotated by two domain experts: a PhD neuro-immunologist and biomedical researcher with 25 years of pharmaceutical and regulatory experience, and a medical doctor with 18 years of clinical practice. Each segment was independently annotated across five dimensions, namely Labovian narrative stage [24], AAO entity spans, emotion, stance, and tone. Disagreements (12.3% of annotations) were resolved through consensus; 47 segments without consensus were excluded. Inter-rater reliability reached Krippendorff’s *α* = 0.78–0.81 across all dimensions [51], exceeding the 0.67 threshold for reliable agreement. The 2,000 annotated segments were drawn from 704 unique threads (mean 2.84 segments per thread) and average 31 words each. These 704 thread-level narratives, each containing at minimum one agent, one action, and one outcome, are the units retained for quadrant analysis.

Corpus 1 carries no independent risk annotation. It supports the independence and prevalence analyses in Sections 6.2 and 6.3, which characterize the joint distribution of the two scores and the share of the corpus that single-axis assessment assigns without regard to modeled harm potential, and it is the corpus on which the operational thresholds were fixed.

#### 5.1.2 Corpus 2: Expert-labeled misinformation benchmark (*N* = 437)

A separate set of Reddit posts was annotated under a binary misinformation protocol for prior work [17], [52], achieving Cohen’s *κ* = 0.82 [53], [54]. The benchmark comprises 437 posts carrying machine-readable labels (310 accurate, 127 misinformation). A further 3 accurate posts carry labels only in free-text form; the encoding difference indicates a separate annotation pass, and they are excluded so that a single protocol defines the evaluation set.

Corpus 2 items are thread-level aggregations of a post title and its full comment set, a different textual unit from the expert-segmented personal accounts that constitute Corpus 1. Their median length is 145 words, with a first quartile of 21 and a maximum of 5,633. Approximately one quarter fall below the length at which a personal experience narrative can be identified, and approximately half exceed the input window of the Phase 1 transformer components. Corpus 2 therefore lies outside the representational scope the Narrative Builder is defined over, and VERITAS scores are not computed on it.

The benchmark instead answers RQ2 directly. Because its labels encode factual accuracy, the question it can settle is what evaluation signal such labels carry: systems trained on those labels and measured against them establish, empirically, how much of the construct is recoverable from annotation of this kind. Since the operational thresholds were fixed on Corpus 1 and play no part in this analysis, no parameter is tuned on the data used to answer RQ2.

### 5.2 Benchmark Evaluation Systems

Six system configurations establish the separability of the Corpus 2 labels. Each is trained or prompted directly on the benchmark’s own text and labels, so none depends on any VERITAS computation, and the strongest among them bounds what those labels can support.

Two term-frequency classifiers (TF-IDF with logistic regression [55]) represent the standard NLP approach to health misinformation classification. Two biomedical-embedding classifiers (frozen BioBERT [13] representations with logistic regression) test whether domain-specific pretraining improves separability. Each is reported in a default and a class-weight-balanced configuration, so that neither a crippled nor an inflated baseline is presented, and all four are evaluated under stratified 5-fold cross-validation, so every post is predicted by a model that never saw it in training.

A zero-shot large language model (claude-sonnet-4-5, temperature 0) tests whether a current-generation model recovers either construct without supervision. It was prompted under two single-word instructions matched to the two axes, one asking whether acting on the post would risk harm and one asking whether the post contradicts medical consensus, with three repetitions per post and majority vote. The paired conditions isolate the effect of the question asked while holding the model, the corpus, and the decision procedure fixed. The same benchmark and systems are also reported in companion work [36].

The class-balanced supervised configurations are the informative cases. A classifier fitted to the benchmark’s own labels and evaluated out of sample establishes an empirical ceiling on the separability those labels afford, and a system assessing a different construct cannot be expected to exceed it. The design therefore tests a structural claim about what these labels encode, not a performance claim about specific model architectures.

### 5.3 Statistical Methods

The association between NTD and NRS is assessed on Corpus 1 through seven statistical tests organized in three families of evidence. The first family measures monotonic association through Pearson correlation, Spearman rank correlation, and Kendall’s *τ*; these are three views of a single underlying relationship rather than three independent confirmations, and they are reported together for robustness to distributional assumptions rather than as accumulating evidence. The second family tests for dependence beyond monotonicity through mutual information [56] and chi-square with Cramér’s *V* [57], either of which could reveal association that the correlation measures would miss. The third establishes what the association supports and whether it could arise by chance, through logistic regression Area Under the Curve (AUC) and distribution-free permutation testing (10,000 iterations). Convergence across the three families, rather than the count of tests, is what the analysis requires. The pre-specified practical-independence criterion is |*r*| < 0.30 for the correlation family and *V* < 0.30 for the categorical measure, corresponding to less than 9% shared variance, below the small-effect-size boundary in Cohen’s conventions [58]; the logistic AUC criterion is < 0.60. Mutual information is estimated with the Kraskov *k*- nearest-neighbor estimator (*k* = 3) and assessed against an empirical permutation null computed on the same corpus rather than against a fixed bit threshold, because *k*-nearest-neighbor estimates carry a sample-size-dependent bias that makes an absolute value non-comparable across corpora. The criterion is one of practical independence rather than orthogonality: what the analysis is required to establish is that the two scores are substantially non-redundant, so that risk is not reliably recoverable from divergence.

On Corpus 2, Youden’s *J* (sensitivity + specificity −1) [59] is the primary metric, computed at each system’s own operating point: the cross-validated class assignment for the supervised systems and the majority-vote flag for the prompted systems. *J* is invariant to class prevalence, which is what makes it preferable to recall or accuracy on an imbalanced benchmark; it is not invariant to the decision threshold, so the operating point at which each value is computed is reported with it. Recall on an imbalanced benchmark rises with flag rate irrespective of discriminative capability, so recall, precision, flag rate, and *F*_1_ are reported alongside *J*, with 95% bootstrap confidence intervals (10,000 resamples of the 437 evaluation posts, seed fixed) for recall and *J*. Threshold-free discrimination (ROC AUC and precision–recall AUC) is not reported because the retained outputs are class assignments and majority-vote flags rather than continuous scores; the comparison is therefore between the operating points the systems actually occupy, and the intervals bound what those operating points can distinguish.

Component contributions to NTD are assessed through ablation against paired *t*-tests with Bonferroni correction, and through weight sensitivity across 11 NTD weight configurations. Both operate on the NTD distribution over Corpus 1 and require no external labels.

## 6 Results

### 6.1 Score Distributions

Table 1 summarizes pipeline production statistics on the 704-narrative corpus.

**Table 1.** VERITAS pipeline production statistics on Corpus 1 (N = 704). The 100% UMLS concept coverage and Krippendorff’s α = 0.78–0.81 inter-rater reliability establish the quality floor of the narrative reconstruction on which Phase 2 scoring depends.

| Metric | Value | Note |
| --- | --- | --- |
| AAO triplets extracted | 4,975 | Mean 7.1 per narrative |
| UMLS concept coverage | 100% | Hybrid normalization (12,101 queries) |
| Clinical quantifier spans | 383 | Identified at BIO tagging |
| Reactive-causation segments | 346 | 346/2,000 segments (17.3%); 137 narratives resolve to reactive causation at thread level |
| Inter-rater reliability | $\alpha = 0.78\text{--}0.81$ | Across all dimensions |

Score distributions across Corpus 1 show mean NTD = 0.393 (SD = 0.196; range 0.002–0.745) and mean NRS = 0.536 (SD = 0.131; range 0.154–0.913). The three NTD pillars reveal characteristic patterns: factual divergence (*D_F_* = 0.358), causal deviation (*D_C_* = 0.421), and evidence calibration (*D_E_* = 0.391). Causal divergence exhibits the highest values, indicating that peer-to-peer health discourse departs from medical consensus most substantially in causal reasoning rather than in terminology.

### 6.2 Dimensional Independence

Five of the seven tests satisfy their pre-specified criteria outright: the three correlation measures, Cramér’s *V*, and the permutation test. Pearson *r* = 0.222 (95% CI [0.150,0.291], *p* < 0.001); Spearman *ρ* = 0.229 (*p* < 0.001); Kendall *τ* = 0.155 (*p* < 0.001); Cramér’s *V* = 0.194 (*χ*^2^ = 26.28, *p* < 0.001); permutation test observed *r* = 0.222 against a null centered on zero (mean −0.000, SD = 0.038, *p* < 0.001). The remaining two qualify the result rather than contradict it. Logistic regression recovers an AUC of 0.617 (SD = 0.027), marginally above the 0.60 criterion, indicating limited rather than absent predictive power. Mutual information *I*(*D*; *R*) = 0.096 bits exceeds its permutation null (null mean 0.014 bits, SD = 0.023), so a nonlinear dependency is detectable; its magnitude nonetheless remains small, consistent with the under-5% shared variance implied by the correlation. Full results are reported in Table 2.

**Table 2.**
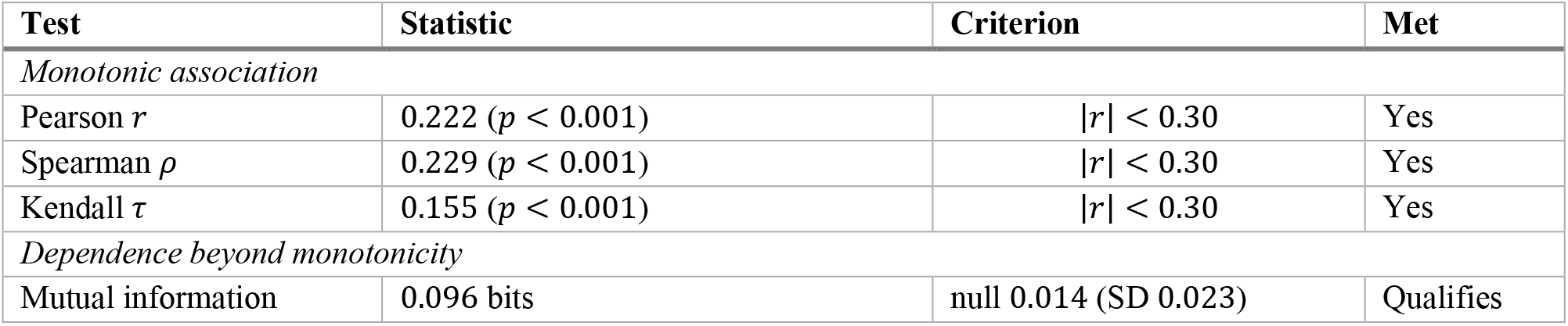
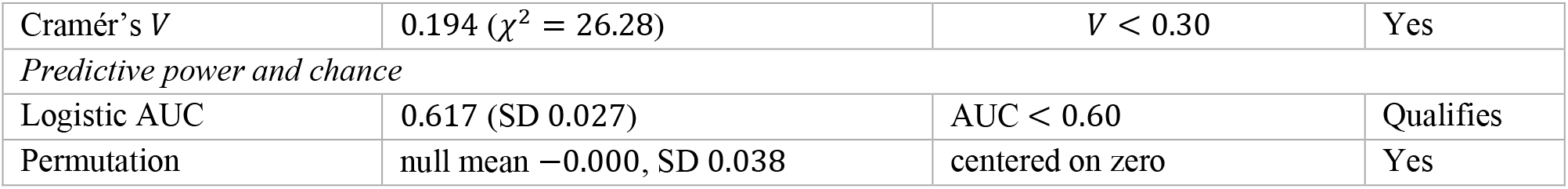
Seven-test analysis of the NTD–NRS association on Corpus 1 (N = 699 classified narratives), organized in three families. The three correlation measures are views of one monotonic relationship. Five tests meet their pre-specified criteria; logistic AUC (0.617, criterion < 0.60) and mutual information (above its per-corpus permutation null) qualify the result, indicating a weak but detectable association below the practical-independence threshold.

Shared variance is *r*^2^ = 0.049, placing 95.1% of the variance in NRS outside what NTD explains. The association is statistically detectable and practically small: it falls below the criterion rather than reaching orthogonality, and the appropriate characterization is a weak association, not independence in the strict sense. What the result supports is the narrower and sufficient claim that the two scores are substantially non-redundant, so risk is not reliably recoverable from divergence and the two dimensions warrant separate measurement.

Figure 3 presents the joint distribution. The local structure is the informative feature: narratives occupying the same NTD band separate widely on NRS, so the failure mode of fact-centric detection is structural rather than incidental.

**Figure 3.**
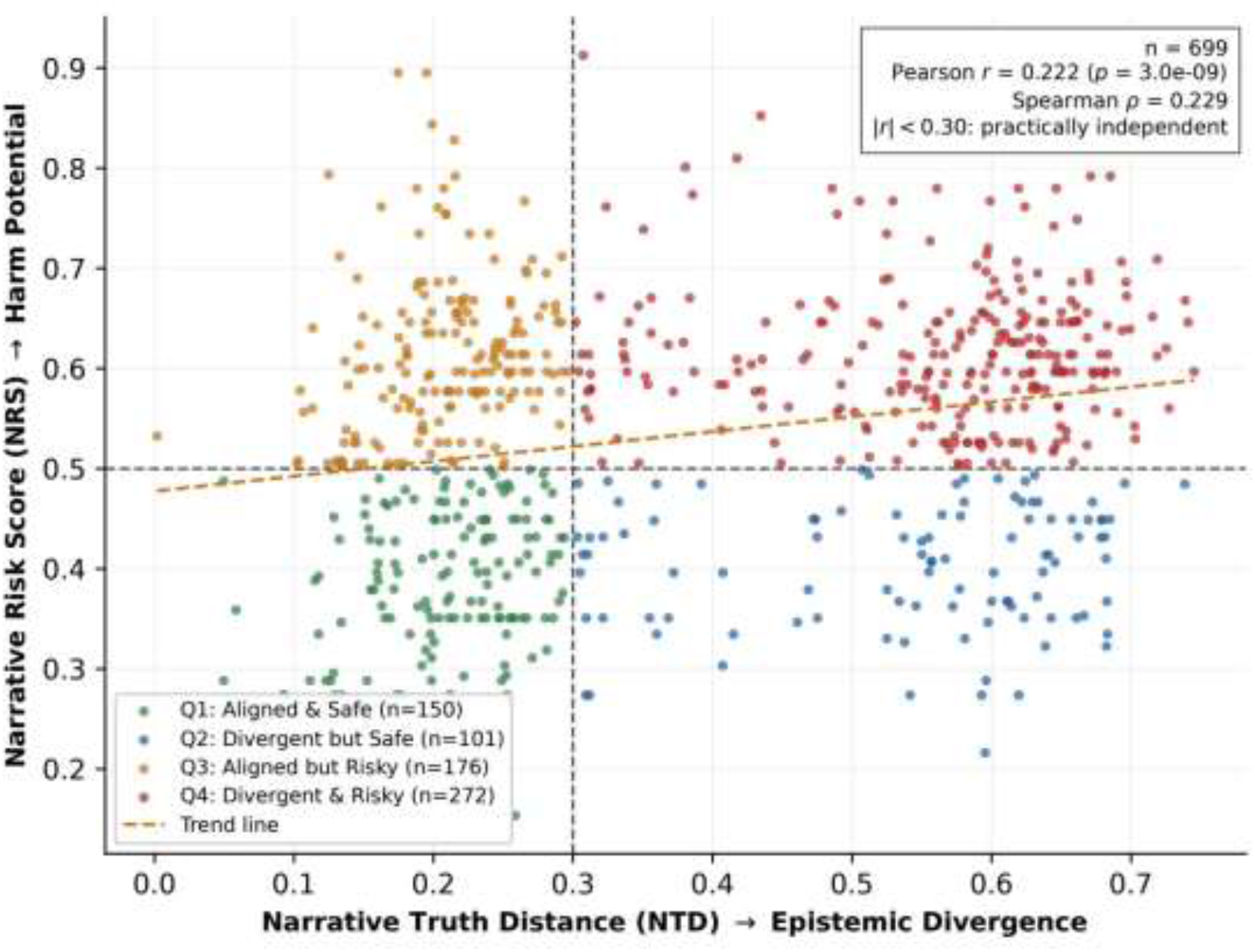
NTD–NRS joint distribution on Corpus 1 (N = 699 classified narratives). Each point is a narrative positioned by its NTD and NRS scores. Dashed lines are operational thresholds (τ_NTD_ = 0.30, τ_NRS_ = 0.50). Pearson r = 0.222; I(D; R) = 0.096 bits. The off-diagonal quadrants (Q2, lower-right; Q3, upper-left) constitute the 39.6% of narratives that single-axis systems mishandle by construction; these cells are defined by the two computed scores, not by an independent reference standard on the risk axis.

**Figure 4.**
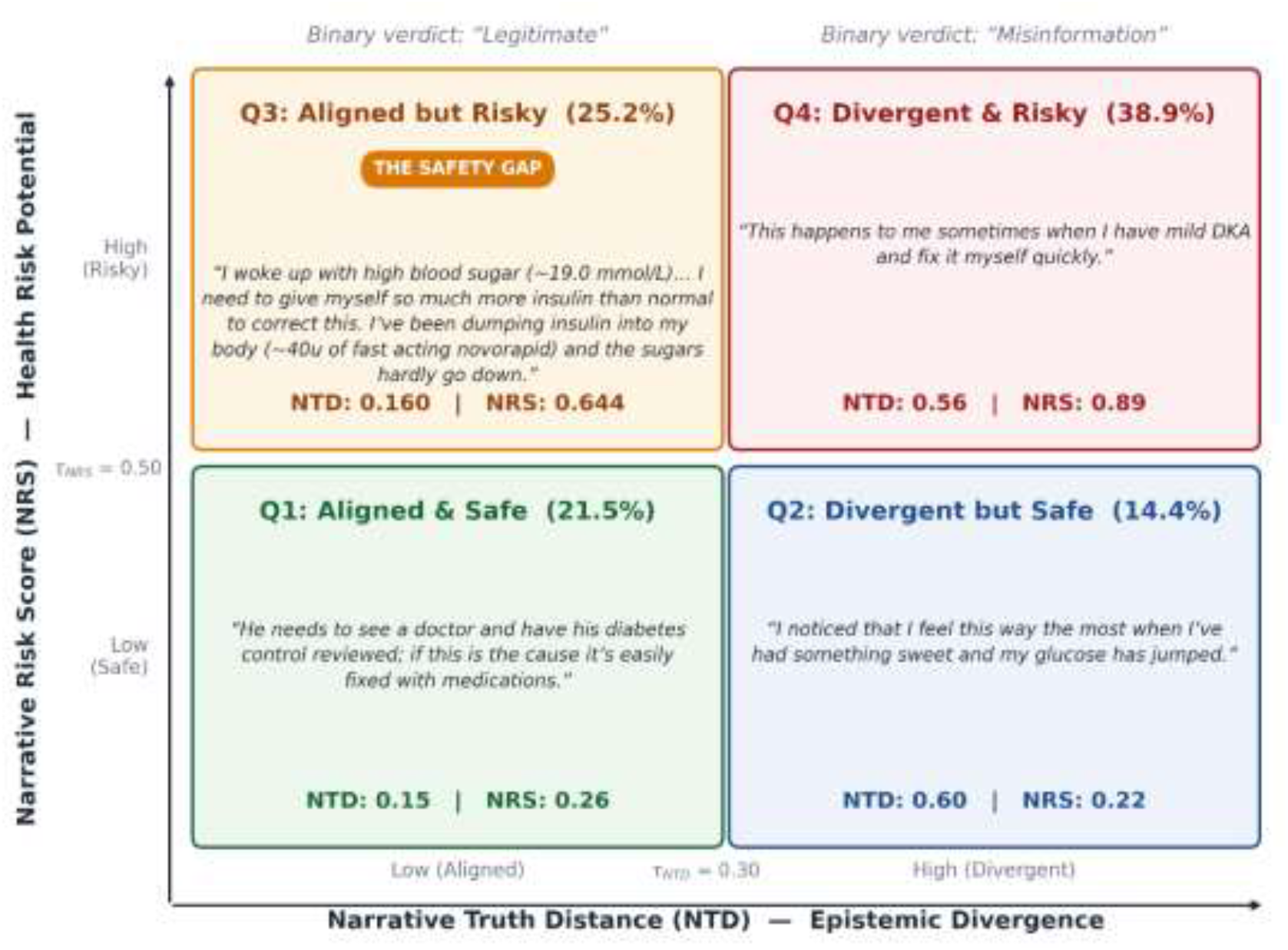
Worked examples of two-dimensional quadrant classification. Each panel presents a representative narrative drawn from the corpus (Q1: narrative 672; Q2: narrative 224; Q3: narrative 41; Q4: narrative 671), lightly condensed for presentation, with its observed NTD and NRS scores. The Q3 panel (Aligned but Risky) illustrates the largest single-axis failure category (25.2%): clinically plausible language describing self-directed management undertaken without any indication of clinical oversight. Q1 and Q4 are the categories where single-axis fact-checking produces the correct outcome; Q2 and Q3 are the categories where it does not.

### 6.3 Quadrant Distribution and Threshold Sensitivity

Table 3 presents the quadrant distribution on Corpus 1.

**Table 3.** Quadrant distribution on Corpus 1 (N = 699 classified narratives; 5 of 704 lacked complete scores). The final column gives the assignment a divergence-only decision rule makes at τ_NTD_ = 0.30 and whether it tracks the modeled harm potential; Q3 (25.2%) is approved and Q2 (14.4%) is flagged without regard to that potential, together the 39.6% share that answers RQ1. Off-diagonal cells are defined by the two computed scores, not by an independent reference standard on the risk axis.

| Quadrant | n | % | NTD | NRS | Divergence-only assignment |
| --- | --- | --- | --- | --- | --- |
| Q1: Aligned & Safe | 150 | 21.5 | 0.207 | 0.384 | Approved; tracks modeled harm potential |
| Q2: Divergent but Safe | 101 | 14.4 | 0.537 | 0.403 | Flagged; does not track |
| Q3: Aligned but Risky | 176 | 25.2 | 0.209 | 0.614 | Approved; does not track |
| Q4: Divergent & Risky | 272 | 38.9 | 0.562 | 0.619 | Flagged; tracks modeled harm potential |
| Q2 + Q3 (off-diagonal) | 277 | 39.6 |  |  |  |

Q3 (Aligned but Risky) accounts for 176/699 narratives (25.2%), with mean NTD of 0.209, below the divergence threshold, and mean NRS of 0.614, above the risk threshold. Q4 (Divergent & Risky) is the largest category at 272/699 (38.9%). Fact-checking and misinformation detection systems classify Q1 (21.5%) and Q4 (38.9%) correctly, together 60.4% of the corpus, and mishandle Q2 and Q3, together 39.6%.

The quadrant pairs sharing an NTD band make the mechanism explicit. Q1 and Q3 differ by 0.002 in mean NTD (0.207 versus 0.209) and by 0.230 in mean NRS (0.384 versus 0.614); Q2 and Q4 differ by 0.025 in mean NTD and by 0.216 in mean NRS. Narratives that a divergence measure cannot tell apart are separated decisively on the risk axis, which is the local expression of the weak global association and the direct answer to RQ1.

Both thresholds were fixed on Corpus 1 before any evaluation. *τ*_NRS_ = 0.50 follows from the NRS construction rather than from a search, as set out in Section 3.3 and specified in Appendix B; in the studied corpus, 19.7% of narratives (138/699) fall within ±0.05 of it, constituting the transition zone. *τ*_NTD_ was set to 0.30 by expert review, to reduce over-flagging of content aligned with traditional health practice, a governance decision reflecting the equity concern that divergence-based suppression disproportionately affects culturally grounded practices (Section 7.4). Robustness was verified through weight sensitivity analysis across 11 NTD pillar configurations (Table 6), which confirms that the practical-independence criterion holds throughout.

### 6.4 Benchmark Separability

RQ2 asks what evaluation signal existing expert-labeled misinformation benchmarks provide for risk-aware assessment. Table 4 sets out how the two corpora differ, and the difference in textual unit is why the question is posed this way: Corpus 2 items are thread-level aggregations rather than the personal health narratives the Narrative Builder is defined over, so VERITAS is not scored on them (Section 5.1). What the benchmark can settle, and does, is how much of the construct its labels afford to any system trained on them.

**Table 4.** Comparison of the two corpora. Corpus 1 supports the independence and prevalence analyses and is where the thresholds were fixed. Corpus 2 supports the separability analysis answering RQ2. No parameter is tuned on Corpus 2.

|  | Corpus 1 | Corpus 2 |
| --- | --- | --- |
| Sample size | $N = 704$ (699 classified) | $N = 437$ |
| Source | 5,386-thread collection | Same collection |
| Textual unit | Expert-segmented narrative segment (mean 31 words) | Title with full comment set (median 145 words) |
| Annotation type | Multi-dimensional (5 dims) | Binary (misinfo/accurate) |
| Protocol | Krippendorff’s $\alpha = 0.78\text{--}0.81$ | Cohen’s $\kappa = 0.82$ |
| VERITAS scored | Yes | No (outside representational scope) |
| Evaluation purpose | Independence, quadrant prevalence, threshold selection | Label separability (RQ2) |
| Threshold tuning | Yes (selection corpus) | Not applicable |

Table 5 reports the six configurations with 95% bootstrap confidence intervals. Recall varies widely, from 8.7% for an unbalanced keyword classifier to 57.5% for class-balanced biomedical embeddings, but recall tracks flag rate throughout, so these differences describe operating points rather than discriminative capability. Discrimination is low across the board: Youden’s *J* of 0.064, 0.349, 0.120, 0.252, 0.201, and 0.168, and no interval reaches 0.45 at its upper bound. With 127 positives the intervals overlap substantially, so the ordering among the four strongest configurations (0.349, 0.252, 0.201, 0.168) is not statistically separated; what the intervals establish is the ceiling, not a ranking.

**Table 5.** Separability of the expert-labeled benchmark Corpus 2 (N = 437; 310 accurate, 127 misinformation). All systems are trained or prompted directly on the benchmark’s own text and labels: supervised systems under stratified 5-fold cross-validation, the zero-shot model under majority vote over three repetitions. The misinformation set (n = 127) is identical across systems, so recall is directly comparable. Flag rate is the proportion of the evaluation set assigned to the positive class; J is Youden’s index (sensitivity + specificity −1), for which 0 denotes chance, computed at each system’s own operating point. Brackets give 95% bootstrap confidence intervals from 10,000 resamples of the 437 posts. The highest J attained is 0.349, by a supervised classifier fitted to the benchmark’s own labels; the intervals overlap, so the ordering among systems is not statistically separated.

| System | Recall [95% CI] | Prec. | Flag rate | $J$ [95% CI] | $F_1$ | Architecture |
| --- | --- | --- | --- | --- | --- | --- |
| TF-IDF + LR (default) | 8.7% [4.0, 13.8] | 61.1% | 4.1% | 0.064 [0.014, 0.119] | 0.152 | Keyword-level |
| TF-IDF + LR (balanced) | 55.9% [47.2, 64.5] | 52.2% | 31.1% | **0.349** [0.252, 0.447] | 0.540 | Keyword-level, class-balanced |
| BioBERT emb. + LR (default) | 25.2% [17.8, 33.0] | 43.8% | 16.7% | 0.120 [0.036, 0.206] | 0.320 | Biomedical embeddings |
| BioBERT emb. + LR (balanced) | 57.5% [48.9, 66.1] | 42.2% | 39.6% | 0.252 [0.152, 0.351] | 0.487 | Biomedical embeddings, class-balanced |
| Zero-shot LLM, risk-focused | 30.7% [22.7, 39.0] | 54.2% | 16.5% | 0.201 [0.114, 0.291] | 0.392 | Prompted, temperature 0 |
| Zero-shot LLM, divergence-focused | 32.3% [24.1, 40.3] | 46.1% | 20.4% | 0.168 [0.078, 0.258] | 0.380 | Prompted, temperature 0 |

Three observations follow. First, the ceiling is low regardless of architecture. The strongest configuration, a classifier fitted to the benchmark’s own labels and evaluated out of sample, reaches *J* = 0.349 with an upper bound of 0.447, and neither frozen biomedical embeddings nor a current-generation language model exceeds it at the point estimate; because the intervals overlap, the claim supported is that no architecture, supervised or prompted, separates these labels well, not that supervision ranks above the alternatives. Second, the paired zero-shot conditions isolate the effect of the question asked while holding model, corpus, and decision procedure fixed. Asking whether acting on a post would risk harm yields *J* = 0.201 [0.114,0.291]; asking whether it contradicts medical consensus yields *J* = 0.168 [0.078,0.258]. The intervals overlap, so querying the risk construct confers no detectable advantage on labels that encode divergence. The near-uniform result therefore reflects the labels rather than the systems.

Third, this is the outcome the independence result predicts. Where epistemic divergence explains 4.9% of the variance in health risk potential, labels annotating factual accuracy cannot carry much signal about behavioral risk, and a modest separability ceiling is expected rather than anomalous. Evaluating risk-aware assessment requires benchmarks annotated for risk. No such benchmark currently exists, which is the evaluation-infrastructure gap identified in Section 7.2.1 and the direct motivation for the risk-annotation program described in Section 7.6. Figure 5 presents the comparison.

**Figure 5.**
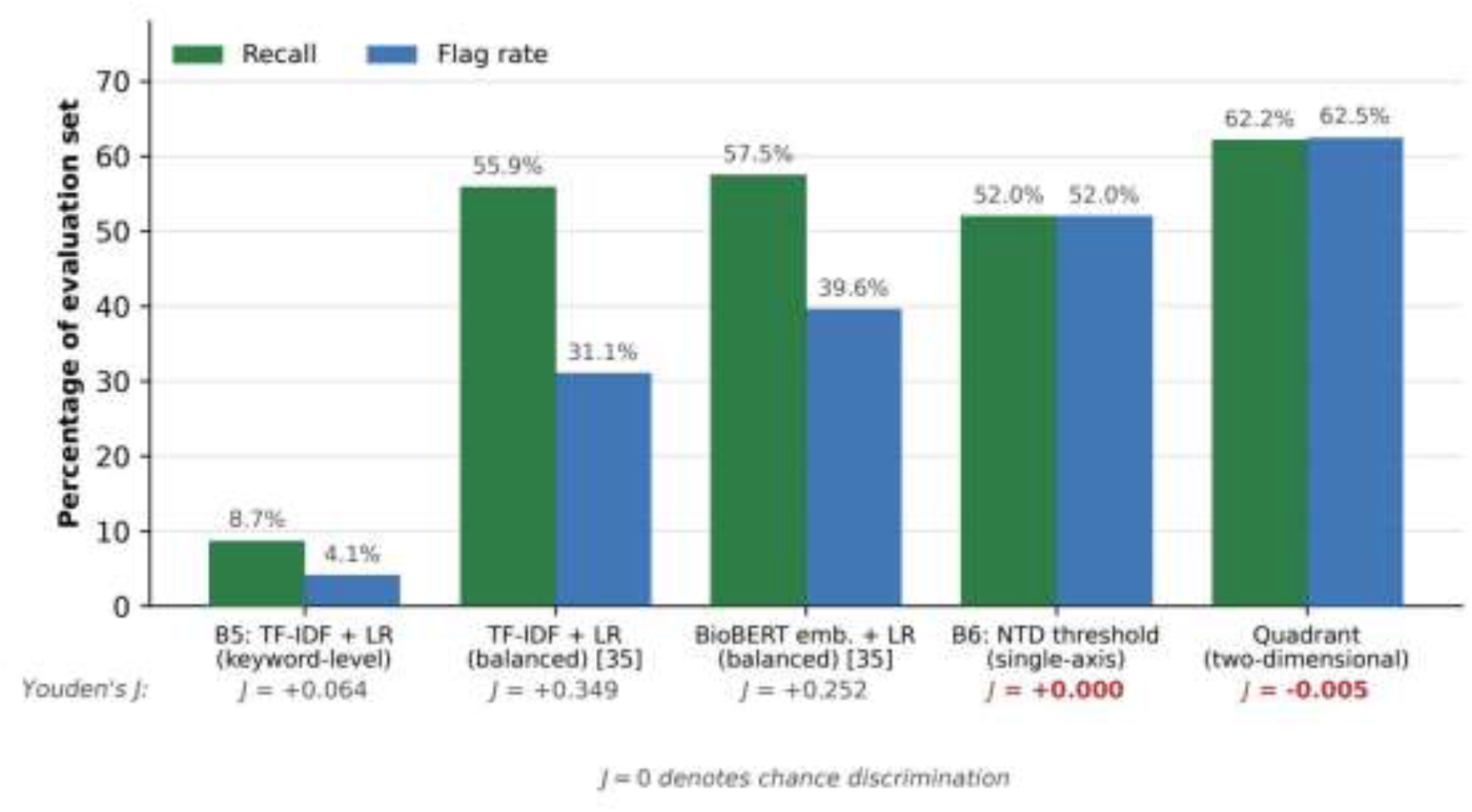
Recall and flag rate for the six configurations evaluated on Corpus 2, each trained or prompted directly on the benchmark’s own text and labels. Recall tracks flag rate throughout, indicating that the differences reflect operating points rather than discriminative capability. Youden’s J is reported beneath each system, where J = 0 denotes chance; the maximum attained is 0.349, by a supervised classifier fitted to the benchmark’s own labels.

A further diagnostic reinforces the point. A majority-class classifier labeling every item “accurate” achieves 70.9% accuracy on Corpus 2 while recovering none of the 127 misinformation instances, exceeding the accuracy of three of the six configurations. Aggregate accuracy on a benchmark with this class balance rewards agreement with the predominant class rather than any capacity to identify potentially harmful content, and is not an informative metric for safety-relevant evaluation.

### 6.5 Component Contributions

Ablation over the NTD pillars identifies causal divergence as the component on which the phenomenon this paper concerns depends. Without *D_C_*, Q3 prevalence falls from 25.2% to 12.4%, whereas removing *D_F_* or *D_E_* leaves it at 28.0% and 27.0% respectively. The shifts in the NTD mean are small relative to its dispersion and are reported as secondary evidence: removing *D_C_* produces the largest shift (*Δ*NTD = −0.0148, *t* = 2.92, *p* = 3.7 × 10^−3^, below the Bonferroni-corrected *α* = 0.0056), removing *D_F_* a smaller shift with a smaller *p* (+0.0117, *p* = 3.2 × 10^−4^), and removing *D_E_* no significant shift (+0.0014, *p* = 0.573). The magnitude of the mean shift is therefore not the basis for identifying *D_C_* as dominant; the collapse of Q3 is. Causal structure is therefore what makes aligned-but-risky content detectable at all; factual terminology and evidence calibration do not substitute for it. Consistent with this, reactive causation is prevalent in the corpus: 346 of 2,000 segments (17.3%) are tagged as outcome-triggering rather than action-causing, and 137 narratives (19.5%) resolve to reactive causation at thread level. Weight sensitivity across 11 configurations maintains the practical-independence criterion throughout, with Q3 prevalence ranging from 12.4% to 28.2% and off-diagonal shares (Q2 + Q3) from 38.2% to 43.6%, as shown in Table 6.

**Table 6.** Weight sensitivity analysis across 11 NTD configurations (7 shown). All configurations maintain the practical-independence criterion (|r| < 0.30), confirming that the core finding is robust to pillar weighting. Fail% denotes the off-diagonal share (Q2 + Q3) that a single-axis rule mishandles by construction.

| Configuration | $w_F/w_C/w_E$ | $r$ | Q3% | Fail% |
| --- | --- | --- | --- | --- |
| Primary | .25/.35/.40 | 0.222 | 25.2 | 39.6 |
| Equal | .33/.33/.34 | 0.209 | 23.9 | 40.2 |
| Causal heavy | .20/.50/.30 | 0.215 | 28.2 | 40.2 |
| Evidence heavy | .20/.30/.50 | 0.234 | 24.5 | 38.8 |
| Extreme $D_C$ | .10/.80/.10 | 0.204 | 28.2 | 39.9 |
| Extreme $D_E$ | .10/.10/.80 | 0.268 | 17.5 | 38.6 |
| No $D_C$ | .50/.00/.50 | 0.131 | 12.4 | 38.2 |

## 7 Discussion

The evidence in Section 6 converges on a single conclusion: the dominant failure mode in online health safety is not the presence of false information but the structural invisibility of accurate content carrying substantial health risk. The Online Health Safety Gap is systematic rather than incidental, affecting 39.6% of the studied corpus.

The first finding concerns how little the two dimensions share. With 4.9% shared variance, knowing how far a narrative departs from consensus says little about whether acting on it would cause harm. Medical fluency is not a proxy for safety. This inverts the foundational premise of health misinformation detection and reframes correctness as an assumption that systematically misfires in peer-to-peer self-care settings. The vocabulary signaling credibility to a detection system is precisely the vocabulary acquired through years of lived experience, self-directed research, or ongoing care, and it is equally available for describing safe and unsafe behaviors.

The second finding follows from the first. The largest mishandled category is not explicitly false narratives but narratives exhibiting *medical-literacy-without-clinical-judgment*: posts by individuals with correct medical vocabulary deploying it, often with confident or pseudo-expert stance, to describe behaviors that may be unsafe without clinical supervision [45], [60]. Detection systems approve this content precisely because it exhibits the epistemic alignment they are designed to detect; it contains nothing for a consensus-alignment detector to notice. Prior qualitative work [2], [3] records the same phenomenon from the user’s perspective: individuals sensed that plausible-looking content could cause harm but lacked any mechanism for separating credibility from safety.

The third finding, and the most instructive for the field, concerns what makes these narratives risky. Ablation identifies causal divergence (*D_C_*) as the dominant NTD component: removing it collapses Q3 prevalence from 25.2% to 12.4%, while removing either of the other two pillars leaves it near 27%. This inverts the expectation of a paradigm that treats misinformation as primarily a terminology problem. The distinction between “my clinician adjusted my metformin” and “I adjusted my metformin myself” turns entirely on narrative structure, namely who initiated the action, under what supervision, and across what temporal sequence, and not on medical vocabulary, which is identical in both formulations. The risk cues that determine safety in peer-to-peer self-care settings are structural features of the whole narrative rather than lexical features of individual claims, and systems operating at the claim level cannot read them irrespective of their terminology handling.

Returning to RQ1, both off-diagonal categories contribute to single-axis mishandling, but asymmetrically and for distinct reasons. Q3, at 25.2% of the corpus, is the mode in which accurate-looking content is approved and evades safety evaluation. Q2, at 14.4%, is the complementary mode in which behaviorally safe but terminologically divergent content would be suppressed by any divergence-triggered moderation rule. The two are not symmetric in frequency, but they are symmetric in what they expose: a single-axis system cannot distinguish either from its diagonal neighbor. Treating Q2 and Q4 as equivalent, or Q3 and Q1 as equivalent, is the structural consequence of projecting the two-dimensional space onto one axis. Both discriminations matter for governance, and the equity dimensions of the Q2 case are taken up in Section 7.4.

Turning to RQ2, the benchmark result places a boundary on what the present evidence can establish. Where a classifier fitted to the benchmark’s own labels reaches *J* = 0.349 (95% CI [0.252,0.447]), and neither biomedical embeddings nor a prompted language model exceeds it at the point estimate, the labels themselves carry limited signal, and no external validation of the risk axis is available from annotation of this kind. This is a constraint on the evaluation infrastructure rather than a property of any system evaluated on it, and it follows directly from the independence result: labels annotating factual accuracy cannot be expected to separate a construct that shares 4.9% of its variance with accuracy. The consequence for the field is set out in Section 7.2.1.

Two implications for deployment follow. The Quadrant is a triage instrument that surfaces content for further review rather than a system making moderation decisions on its own, and the continuous NTD and NRS scores are preserved throughout the pipeline so that the operating point remains a deployment choice. Where that point should sit is an empirical question that a risk-annotated benchmark would settle and the present evidence cannot: establishing the precision of the risk axis requires labels that encode risk. Section 7.6 identifies this as the priority next step.

### 7.1 Objective Misspecification, Not Inadequate Modeling

The 39.6% off-diagonal share under single-axis classification is not anomalous; it is consistent with a pattern documented across the health NLP literature. Prior work reports 12% recall for a Term Frequency–Inverse Document Frequency (TF-IDF) NLP health misinformation classifier [17], 34% recall for GPT-4o on underrepresented mental health categories [18], F1- scores as low as 0.355 across 33 cancer-information extraction studies [19], and 33% F1 collapse when clinical Named Entity Recognition (NER) models trained on synthetic data are applied to real clinical text [61]. These deficits persist across architectural generations, the signature of a structural cause rather than model-specific shortcomings. We term this *objective misspecification*: existing systems are optimized for truth-conditional classification against medical consensus, but the operational requirement in peer-to-peer self-care settings is risk-aware assessment of whether a described behavior is safe to act on under uncertainty. These are not variants of the same task, and improvements within the former will not resolve the deficit in the latter.

The misspecification extends to measurement. The majority-class result reported in Section 6.4 shows that agreement with the predominant class can outscore every system that actually recovers safety-critical content. Evaluating safety-relevant systems on aggregate accuracy is itself a categorical error, of the same kind as the task misspecification it is measuring.

The same lens extends to the guardrail systems of large language models. Hallucination-focused guardrails [41], [42] do not intervene when a model produces consensus-consistent output encoding an unsafe action pattern, because no factual error has occurred. The two-dimensional requirement established here applies directly: safety-aware LLM guardrails in health contexts require independent harm assessment alongside epistemic grounding.

### 7.2 What Must Change: Four Governance Obligations

The evidence reported here is not descriptive of a problem; it is prescriptive of a structural response. Four classes of actors in the online health information environment carry specific obligations that follow directly from the findings.

#### 7.2.1 Benchmark Design

Existing health misinformation benchmarks are misspecified by construction. By annotating content for factual accuracy alone, they reward systems for detecting Q4 (38.9% of the corpus) while providing no evaluation signal for Q3 (25.2%). A system that perfectly detects Q4 and ignores Q3 entirely would score highly on current benchmarks while offering almost no protection against the dominant failure mode in peer-to-peer health discourse. The separability ceiling reported in Section 6.4 quantifies the consequence: on labels of this construction, the best any system achieved is *J* = 0.349, so the benchmark cannot distinguish a risk-aware system from a divergence-based one whatever either does. Future benchmark construction must therefore pair factual accuracy labels with independent behavioral risk annotation, applied over AAO-structured narrative representation so that both dimensions are scored. This replaces current evaluation infrastructure rather than extending it.

#### 7.2.2 Moderation Architecture

Platform moderation pipelines implementing binary accept-or-reject decisions for health content are architecturally incapable of the proportionate interventions that governance frameworks increasingly require. The EU AI Act [39] mandates that intervention severity be commensurate with assessed risk; single-axis systems cannot operationalize this because they cannot distinguish Q2 (divergent but safe) from Q4 (divergent and risky), nor Q3 (aligned but risky) from Q1 (aligned and safe). Two-dimensional classification makes these discriminations structurally available. Q3 narratives, constituting 25.2% of the studied corpus, warrant targeted safety advisories rather than removal: the content is consensus-aligned, so removal sacrifices genuine informational value, while the harm is conditional on action and a warning addresses that conditional directly. Q2 content warrants contextual annotation rather than suppression, since divergence-based suppression applied to Q2 disproportionately restricts culturally grounded health practices that are safe but terminologically distant from Western biomedical orthodoxy [46]. Platforms that wish to implement proportionate health governance must replace accept-or-reject architectures with systems that compute divergence and risk on independent pathways.

#### 7.2.3 LLM Guardrail Objectives

Large language models mediating health information through chat interfaces, search summarization, and embedded assistants [40], [41], [42], [62] require guardrails that assess what is said and what would follow if a user acts on it. Hallucination detection, evaluating factual consistency against a knowledge base, is necessary but not sufficient: it inherits the epistemic- only objective that limits binary classifiers and does not intervene when a model generates consensus-accurate text encoding an unsafe behavioral pattern. The zero-shot results in Section 6.4 bear on this directly. Prompting a current-generation model to assess harm rather than accuracy did not improve separability, which indicates that instruction alone does not substitute for an architecture that scores the two constructs separately. Independent harm assessment is a prerequisite for safety-aware guardrails in any AI system that generates, curates, or surfaces health information in non-clinical settings, not an optional enhancement over hallucination detection.

#### 7.2.4 Clinical Versus Non-Clinical Risk Framing

Risk stratification in clinical settings feeds into institutional workflows with professional oversight: a clinician interprets the output, a protocol governs the response, and a regulated pathway enforces proportionate action. None of these conditions holds in the peer-to-peer information environment. There is no clinician to interpret the Quadrant’s output on the user’s behalf, no protocol to govern the platform’s response automatically, and no regulated pathway to enforce action. Non-clinical risk-aware assessment must therefore be designed around the requirement that an individual user, not a trained professional, can evaluate the relevance of the assessed risk to their own circumstances and decide whether to seek further guidance. This is why the evidence-based explanation accompanying each quadrant assignment is a structural requirement: without it, a label communicates a verdict without the reasoning that allows a user to assess whether that verdict applies to them. Figure 6 illustrates this for a Q3 narrative, showing how the two scores and their constituent signals are surfaced alongside the classification. The shift from clinical to non-clinical risk-aware assessment is a shift from a supervised to an unsupervised environment, and the design obligations change accordingly.

**Figure 6.**
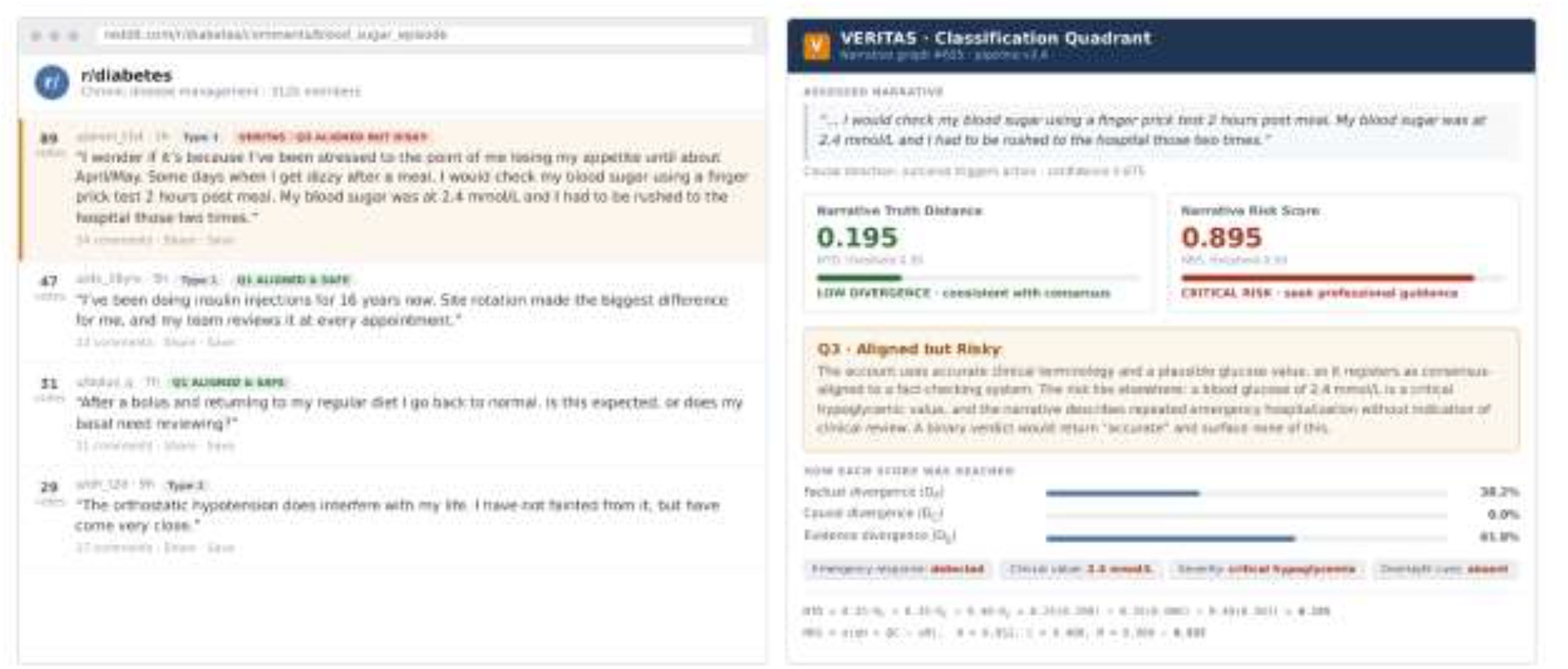
Illustration of the browser interface of the Classification Quadrant, shown for narrative graph #655. The source thread appears on the left and the assessment on the right. The display separates the low-divergence assessment (NTD = 0.195, below the 0.30 threshold: the narrative is factually consistent with medical consensus) from the elevated health risk assessment (NRS = 0.895, above the 0.50 threshold: the behavior described carries significant risk if acted upon without clinical supervision), together with the specific narrative elements driving each score. A binary label would show only “accurate,” leaving the health risk invisible to the user.

### 7.3 Theoretical Implications

The two-dimensional reframing carries weight beyond online health safety. The dominant architectural pattern in safety-critical AI, that is, truth-alignment in health, factual consistency in LLMs, and policy-compliance in content moderation, is a single-axis objective derived from the domain’s epistemic standard. The results presented here indicate that where users make consequential decisions under uncertainty, a fact-based epistemic objective is structurally inadequate regardless of how well it is optimized. The pattern the Quadrant instantiates, namely independent scoring of epistemic alignment and behavioral consequence combined through a partition that preserves both dimensions as actionable, is applicable to any information domain in which credibility and safety can diverge, including financial advice, legal guidance, and safety-relevant technical information. The present work demonstrates the pattern in one domain; testing its generality across domains is work the field should undertake.

### 7.4 Health Equity and Epistemic Justice

The weak association between the two dimensions (*r* = 0.222, 4.9% shared variance) carries an equity implication extending beyond the technical finding. Because divergence carries little information about behavioral safety, a moderation regime treating divergence as a proxy for harm mishandles 39.6% of content: it suppresses Q2 content (14.4% of the corpus) that is divergent but behaviorally safe, while leaving Q3 content (25.2%) that is aligned but behaviorally risky without any safety signal. Divergence-based suppression applied to Q2 disproportionately affects culturally grounded health practices that are terminologically distant from Western biomedical consensus but behaviorally safe, restricting the informational access of populations relying on community-validated traditional approaches without any corresponding safety benefit [2], [3].

Reframing content verification from “does this diverge from consensus?” to “does this carry elevated harm potential?” operationalizes a distinction health equity requires. Biomedical consensus is not culturally neutral; it reflects particular research priorities, pharmaceutical investment patterns, and the healthcare infrastructure of high-income countries. Alternative health systems such as traditional Chinese and African medicine, Ayurvedic practice, Indigenous healing, and folk remedies represent distinct epistemological foundations, not incorrect versions of biomedicine. The Classification Quadrant acknowledges this computationally: Q2 is the category in which divergence from biomedical orthodoxy coexists with low harm potential, and its separate identification enables a moderation response proportionate to actual risk, namely contextualization rather than suppression. Treating Q2 and Q4 as equivalent, as single-axis divergence detection necessarily does, reproduces the epistemic hierarchy at the level of automated governance.

### 7.5 Limitations

The evidence establishes the prevalence and structure of the Online Health Safety Gap. It does not establish the precision of the risk axis, because the available expert-labeled benchmark encodes factual accuracy rather than behavioral risk, and its items are thread-level aggregations rather than the personal health narratives the Narrative Builder is defined over. Direct validation requires a benchmark annotated for risk, described in Section 7.6. Until one exists, the framework supports triaging content for further review rather than unsupervised moderation.

The prevalence findings answering RQ1 are computed from the NRS and are therefore dependent on it: the 25.2% Q3 share and the 39.6% off-diagonal share describe how two separately computed scores partition the corpus, not an independently verified count of harmful narratives. RQ2 does not inherit this dependency, since the separability ceiling is established by classifiers trained on the benchmark’s own labels with no VERITAS score involved.

The severity component of NRS draws on SNOMED CT [15] and the evidence pillar on PubMed, so the *H* component of Equation [eq:nrs] may overstate the risk of practices grounded in non-Western medical traditions, that is, assign them higher harm potential than their behavioral profile warrants, when constructing the risk vectors and not only when evaluating specific content. The architectural separation of NTD and NRS (ℱ_NTD_ ∩ ℱ_NRS_ = Ø) confines this to specific NRS components; the two-dimensional framework is knowledge-base-agnostic, and the WHO ICD-11 Traditional Medicine module, the TICM ontology, and the Traditional Knowledge Digital Library are available substitutions. NRS also models risk from textual cues alone, so user-level factors such as comorbidities and medication history would enable personalized rather than population-average assessment.

Q3 prevalence was measured in a corpus dominated by chronic disease self-management. The corpus was constructed around these conditions because prior qualitative work identified them as the health topics most commonly sought online [2], [3], not because they were expected to yield a particular quadrant distribution. Whether aligned-but-risky content is more or less prevalent in other settings is untested: the 25.2% figure characterizes the corpus studied, and acute care, mental health, reproductive health, and pediatric contexts warrant separate measurement before any general claim about peer-to-peer health discourse can be made.

### 7.6 Future Work

Five directions extend the present results. The priority is a benchmark annotated for behavioral risk alongside factual accuracy, using AAO-structured narrative representation, with inter-rater reliability reported for the risk dimension; this would close the evaluation-infrastructure gap identified in Section 7.2.1 and is a precondition for measuring the precision of the risk axis at all.

Applying the two-dimensional framework at full corpus scale and across platforms and languages would test whether the aligned-but-risky prevalence is a general property of peer-to-peer health discourse or a feature of specific platform conventions. Cross-domain validation across health domains with distinct behavioral-risk profiles, namely mental health, reproductive health, pediatric care, and acute presentations, would test the Quadrant’s generality beyond the chronic-disease self-management context studied here. Evaluation in deployment settings, including platform moderation pipelines, LLM guardrail integrations, and seeker-facing interfaces, would test whether the theoretical benefits translate into practical safety improvements, including the question of whether users shown two-dimensional assessments make more informed decisions than users given a single-label verdict. Finally, the zero-shot comparison reported in Section 6.4 should be extended across prompting strategies, few-shot and chain-of-thought conditions, and additional model families, to establish whether prompted large language models can approximate two-dimensional assessment without the explicit architectural separation used here. Such a comparison tests model capability rather than dimensional adequacy, and requires careful experimental design to isolate risk assessment from general language understanding [63].

## 8 Conclusion

This paper identifies a structural failure in how online health information is evaluated. Factual accuracy is not a reliable indicator of safety in peer-to-peer health discourse: content that aligns with medical consensus can describe behaviors that are harmful when enacted without clinical supervision, and current systems are not designed to detect that distinction. Across 699 classified narratives, epistemic divergence leaves 95.1% of the variance in health risk unexplained, 39.6% of content falls in quadrants that single-axis systems mishandle by construction, and the dominant discriminating signal is causal structure rather than medical vocabulary.

The evaluation infrastructure carries the same limitation. On an expert-labeled misinformation benchmark, the highest discrimination attained by any system is Youden’s *J* = 0.349 (95% CI [0.252,0.447]), reached by a supervised classifier fitted to those labels directly, and neither biomedical embeddings nor a prompted large language model exceeds it at the point estimate.

Labels annotating factual accuracy cannot separate a construct that shares 4.9% of its variance with accuracy, so the gap identified here is at present measurable in prevalence but not in detection performance.

For an individual user, the implication is direct: credible-looking health information is not necessarily safe to follow. The ability to distinguish what is accurate from what is safe to act on in one’s own circumstances becomes essential in environments where decisions are made without professional guidance. The Classification Quadrant operationalizes that distinction computationally, providing both an assessment and the evidence behind it.

These findings call for changed objectives, not better models. Four obligations follow. Benchmark designers must pair accuracy annotation with independent behavioral risk annotation; current benchmarks reward detection of Q4 (38.9% of the corpus) while providing no signal for Q3 (25.2%) or Q2 (14.4%), and the separability ceiling reported here quantifies what that costs. Platform architects should replace binary-logic moderation pipelines with two-dimensional systems capable of proportionate, risk-aware responses; accept-or-reject architectures cannot make the discriminations that modern governance frameworks require. LLM guardrail designers must incorporate independent harm assessment alongside hallucination detection; consensus-aligned output encoding an unsafe behavioral pattern is not a hallucination, and hallucination-focused guardrails will not detect it. Non-clinical implementations must treat evidence-based explanation as a structural requirement rather than an interface feature, because without it a categorical label gives a user no basis to judge whether the assessed risk applies to their own circumstances.

The conceptual infrastructure for distinguishing credibility from safety is available; the measurement infrastructure is not. The Quadrant functions as a triage instrument that surfaces content for further review, and calibrating it for any deployment context requires a benchmark annotated for behavioral risk, which does not yet exist. Building that benchmark is the precondition for the work that follows: evaluation and extension across the information environments where individuals make consequential health decisions without clinical guidance.

## Data Availability

The Reddit corpus used in this study cannot be redistributed due to Reddits Terms of Service and user privacy considerations; the data were collected via the official Reddit API (PRAW) in compliance with platform terms and established ethical guidelines for internet research. The VERITAS implementation producing the NTD and NRS scores is described and released with the companion prior-work publications. The biomedical knowledge bases used (UMLS, SNOMED CT, SemMedDB, PubMed) are publicly available under their respective licensing terms.

## Appendix A: NTD Weight Derivation

The NTD component weights (*w_F_* = 0.25, *w_C_* = 0.35, *w_E_* = 0.40) were determined through a two-stage process combining theoretical grounding with empirical optimization.

### Stage 1: Theory-driven bounds

Exploratory analysis of the Reddit health corpus revealed distinct error patterns: 30% of narratives contained terminology misuse, 75% contained causal oversimplification, and 85% contained unsourced claims. These frequencies established approximate weight ranges prioritizing the most prevalent error types: *w_F_* ∈ [0.20,0.30] (terminology, the least frequent), *w_C_* ∈ [0.30,0.40] (causality, directly harmful through faulty reasoning), and *w_E_* ∈ [0.35,0.45] (evidence, the most frequent and most harmful through overconfident advice).

### Stage 2: Empirical optimization

Within these ranges, grid search (step size 0.05) identified the weights (0.25, 0.35, 0.40) that maximized discriminative power across the observed NTD range [0.002, 0.745] while maintaining the practical-independence criterion (|*r*| < 0.30) across all 11 configurations tested. Ablation supports the ordering: removing *D_C_* produces the largest shift in the NTD distribution and the largest change in Q3 prevalence of the three pillars (Section 6.5).

## Appendix B: NRS Component Specification

The full expanded NRS formula is:

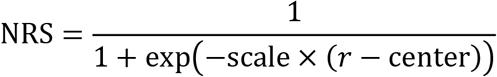

where *r* = *α* ⋅ *H* + *β* ⋅ *C* − *γ* ⋅ *M* and the parameters are specified in Table 7. The centering constant maps a raw risk of 0.75, the midpoint of the raw-risk scale, to NRS = 0.50. The transformation is monotone and preserves the ordering of raw risk scores; its role is to map an unbounded linear combination onto the [0,1] interval with a graded transition band rather than a categorical boundary. On the classified corpus the raw-risk median is 0.790, so NRS = 0.50 falls at approximately the 36th percentile of the score distribution rather than at its median; at center = 0.80 the median NRS would be 0.486, the setting at which *τ*_NRS_ = 0.50 would coincide with the empirical median. This construction is the basis for the operational threshold *τ*_NRS_ = 0.50 used in Section 6.3.

**Table 7.**
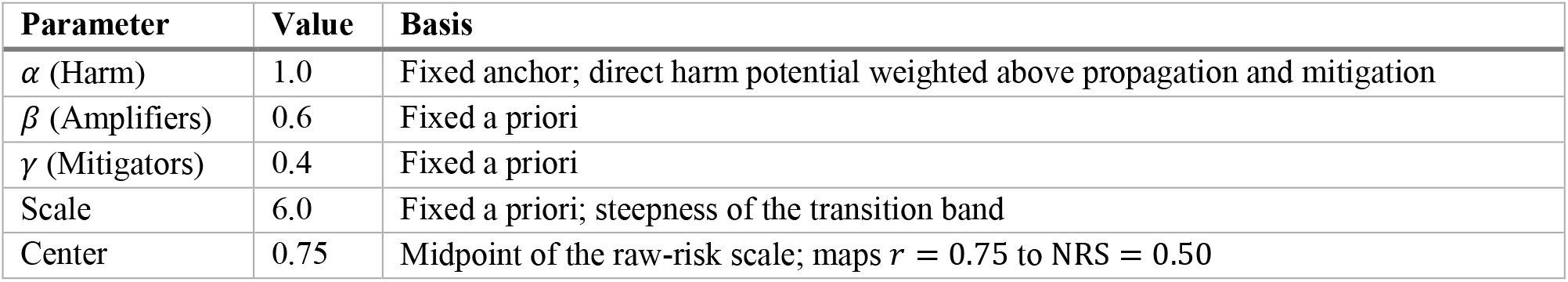
NRS parameters and the basis on which each was set. All five are fixed constants specified before the canonical pipeline run; none is estimated from data or tuned against any label set.

| Parameter | Value | Basis |
| --- | --- | --- |
| $\alpha$ (Harm) | 1.0 | Fixed anchor; direct harm potential weighted above propagation and mitigation |
| $\beta$ (Amplifiers) | 0.6 | Fixed a priori |
| $\gamma$ (Mitigators) | 0.4 | Fixed a priori |
| Scale | 6.0 | Fixed a priori; steepness of the transition band |
| Center | 0.75 | Midpoint of the raw-risk scale; maps $r = 0.75$ to $\text{NRS} = 0.50$ |

*Harm potential* (*H*) uses geometric aggregation: *H* = (*H*_severity_ × *H*_immediacy_ × *H*_vulnerability_)^1/3^. The geometric mean requires all three dimensions to be elevated for high composite harm, preventing false positives from a single extreme component. *H*_severity_ maps outcomes to SNOMED CT clinical finding severity (no harm = 0.0; mild = 0.2; moderate = 0.4; severe = 0.6; hospitalization = 0.8; death or organ failure = 1.0). *H*_immediacy_ captures temporal urgency (immediate = 0.95; days = 0.80; weeks = 0.60; months = 0.40; years = 0.15). *H*_vulnerability_ assesses population risk factors (terminal or immunocompromised = 0.90; elderly, pregnant, or children = 0.80; chronic condition = 0.75; general population = 0.50; healthy adults = 0.45).

*Contextual amplifiers* (*C*) aggregate reach and influence factors: *C* = 0.4 ⋅ *C*_virality_ + 0.4 ⋅ *C*_certainty_ + 0.2 ⋅ *C*_emotional_. Equal emphasis on virality and certainty mismatch reflects their comparable influence on behavioral adoption; the lower weight on emotional intensity acknowledges that emotional framing can be appropriate in health contexts. *Protective mitigators* (*M*) quantify protective language: *M* = 0.4 ⋅ *M*_disclaimer_ + 0.4 ⋅ *M*_consult_ + 0.2 ⋅ *M*_hedging_. Explicit protective measures, namely disclaimers and consultation recommendations, receive equal emphasis at 40% each; hedging receives 20% because it is easily employed without substantive protective intent.

*Parameter provenance and scope of validation.* The parameters in Table 7, and the sub- component weights of *C* and *M*, were not estimated from data. They are fixed constants specified before the canonical pipeline run and documented in the released configuration; the weights do not sum to 1, reflecting the distinct measurement scales of the three components and the priority accorded to direct harm potential over propagation and mitigation. No logistic regression, maximum likelihood, or grid search over a labeled target was performed, so there is no dependent variable, annotator, or fitting sample to report. No component of the NRS reads the expert misinformation labels of Corpus 2, and no parameter is tuned against any outcome label on Corpus 1, which carries no independent risk annotation. The NRS is therefore neither a criterion-validated risk model nor an internally optimized construct. It is a theory-specified construct, and the validation claimed for it in this paper is delimited to construct-level evidence: the derivation of its components from the Kaplan-Garrick formalism [22], the architectural separation of its inputs from those of NTD, and sensitivity analysis across its parameterization. Across scale ∈ {4.0,6.0,8.0} and center ∈ {0.70,0.75,0.80}, the NTD–NRS correlation remains within *r* = 0.216 to 0.231 and the Q3 share ranges from 17.7% to 33.0%, so the practical- independence result does not depend on the particular sigmoid parameterization chosen. Convergent validation against independent expert ratings of harm potential is not part of this evaluation and is set out in Section 7.6.

## Appendix C: Narrative Builder Architecture and Validation

This appendix summarizes the Phase 1 architecture and its validation so that the present paper can be assessed without reference to [35].

### C.1 Architecture

Phase 1 applies two transformers to segmented narrative text. A DistilBERT encoder (66M parameters) with four classification heads assigns Labovian narrative stage, emotion, stance, and tone, with task loss weights of 1.0, 0.8, 0.8, and 0.8 respectively, assigned manually to modestly upweight the primary Labovian task. A BioBERT encoder (110M parameters) with a conditional random field layer performs BIO sequence tagging for agent, action, and outcome spans, using first-subword masking and CRF negative log-likelihood.

### C.2 Data partitioning

Two domain experts, a biomedical researcher with 25 years of pharmaceutical and regulatory experience and a medical doctor with 18 years of clinical practice, independently annotated 2,000 narrative segments drawn from 704 threads (mean 2.84 segments per thread), reaching Krippendorff’s *α* = 0.78–0.81 across dimensions; 12.3% of annotations required consensus adjudication.

Both models partition on the thread identifier, so segments originating in the same thread never appear in different partitions, and each carves a held-out test set (15% of threads) before five-fold grouped cross-validation on the remaining development pool. The two partitions are drawn independently: the span extractor is tested on 286 segments from 100 unseen threads, and the interpretive classifier on 282 segments from 101 unseen threads. Zero thread overlap across train, validation, and test partitions, and across all five folds, was verified programmatically against the production splitting code.

### C.3 Interpretive classification

Table 8 reports per-task macro-F1 across the five grouped folds. Reported standard deviations are population standard deviations across folds.

**Table 8.**
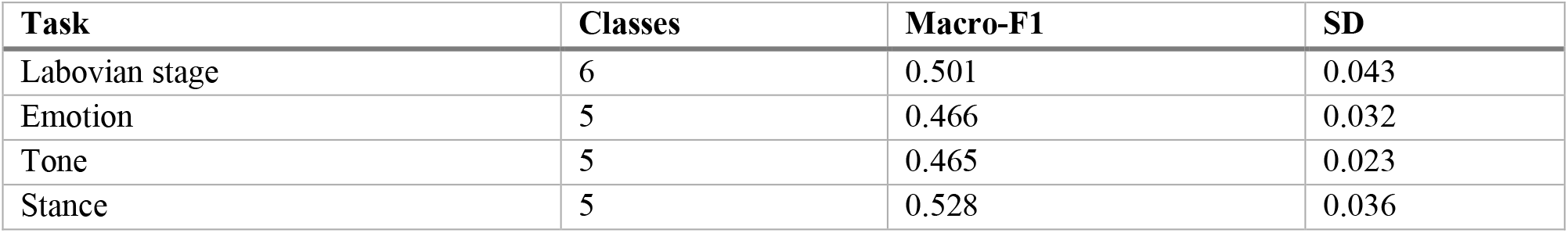
Interpretive classification performance, five-fold grouped cross-validation.

| Task | Classes | Macro-F1 | SD |
| --- | --- | --- | --- |
| Labovian stage | 6 | 0.501 | 0.043 |
| Emotion | 5 | 0.466 | 0.032 |
| Tone | 5 | 0.465 | 0.023 |
| Stance | 5 | 0.528 | 0.036 |

Macro-F1 is depressed by low-support minority classes rather than by uniform weakness. Averaged across the five folds, per-class F1 on the primary Labovian task is 0.735 (Orientation), 0.599 (Evaluation), 0.560 (Complicating Action), 0.533 (Abstract), 0.320 (Coda), and 0.259 (Resolution). The weakest classes across the remaining tasks are Anger (0.071), Skeptical (0.074), and Questioning (0.209), with 54, 73, and 98 instances respectively in the cross-validation pool. These dimensions contribute contextual signal to the NRS amplifier and mitigator components rather than serving as standalone classifications, so their minority-class performance does not propagate directly to the quadrant assignment.

### C.4 AAO span extraction

Table 9 reports held-out span-level performance under strict IOB2 seqeval matching, the convention used throughout the paper. Cross-validated span-F1 is 0.851 ± 0.007.

**Table 9.** AAO span extraction on the held-out test set (286 segments, 100 unseen threads, 2,138 gold spans by first-token count), strict IOB2.

| Entity type | Precision | Recall | $F_1$ |
| --- | --- | --- | --- |
| ACTION | 0.945 | 0.857 | 0.899 |
| AGENT | 0.880 | 0.789 | 0.832 |
| OUTCOME | 0.774 | 0.751 | 0.763 |
| Micro average | 0.883 | 0.813 | 0.846 |
| Macro average | 0.867 | 0.799 | 0.831 |

Under the same convention and on the same held-out set, a TF-IDF and logistic regression tagger reaches span-F1 0.773 and BioBERT without the CRF layer reaches 0.838, against 0.846 for the full BioBERT+CRF configuration. McNemar tests with continuity correction at the entity-span level give *χ*^2^ = 288.69 for TF-IDF against BioBERT without CRF and *χ*^2^ = 300.27 against BioBERT+CRF (both *p* < 0.001), while the difference between the two transformer configurations is not significant (*χ*^2^ = 1.23, *p* = 0.266). The CRF layer therefore contributes structural validity to the tag sequence rather than a significant gain in span-level accuracy.

### C.5 Concept normalization

Extracted entities are normalized against UMLS through a three-stage cascade. Across 12,101 normalization queries, coverage is complete: 7,769 (64.2%) resolved by exact match, 3,185 (26.3%) by approximate string matching, and 1,147 (9.5%) by BioBERT semantic similarity, the last at a mean cosine similarity of 0.9499.

### C.6 Production statistics

Applied to the 704 annotated threads, Phase 1 produced 4,975 AAO triplets (mean 7.1 per narrative) and identified 383 clinical quantifier spans at the BIO tagging stage. Reactive causation, in which an outcome triggers the action rather than following from it, was detected in 346 of 2,000 segments (17.3%); at thread level, 137 narratives (19.5%) resolve to reactive causation. Quantifier counts reported elsewhere in this paper refer to the BIO tagging stage.

## Data and Code Availability

The Reddit corpus used in this study cannot be redistributed due to Reddit’s Terms of Service and user privacy considerations; the data were collected via the official Reddit API (PRAW) in compliance with platform terms and established ethical guidelines for internet research [50]. The VERITAS implementation producing the NTD and NRS scores is described and released with the companion prior-work publications [35], [36]. The biomedical knowledge bases used (UMLS, SNOMED CT, SemMedDB, PubMed) are publicly available under their respective licensing terms.

## Acknowledgment

The authors acknowledge the assistance of the clinician and biomedical researcher who annotated the dataset, whose work was crucial to the success of this project.

## AI Use

Portions of this manuscript, specifically prose revisions in the Introduction, Discussion, and Conclusion sections, were edited and streamlined for length and clarity using a generative language model (Claude, Anthropic) [64]. AI assistance was limited to prose revision and structural reorganization to meet journal formatting requirements; it was not used for data analysis, statistical computation, or interpretation of results. All scientific content, analyses, and interpretations were authored and verified by the human authors, who reviewed and approved all AI-assisted edits and take full responsibility for the content.

